# Modifiable Contributors to Socioeconomic Inequality in Brain Aging

**DOI:** 10.64898/2026.08.13.26360373

**Authors:** Harry W. Richardson, Austin J. Dibble, Connor Dalby, Katie A. Robertson, Frederick K Ho, Donald M. Lyall, Monika Harvey, Michele Svanera

## Abstract

**Importance:** Socioeconomic disadvantage is associated with accelerated brain aging. However, the modifiable factors accounting for this association, and whether they differ across socioeconomic indicators, remains unclear.

**Objective:** To determine whether modifiable risk factors account for socioeconomic differences in the brain age gap, and whether their contributions differ between individual-level and area-level socioeconomic indicators.

**Design, Setting, and Participants:** This cohort study used data from the UK Biobank, a population-based cohort recruited at ages 40 to 70 years from 2006 to 2010. Participants with T1-weighted and T2-FLAIR brain MRI at the first imaging visit were eligible; 7700 used for model development in previous work were excluded, yielding 36 878. Data were analyzed from April to July 2026.

**Exposures:** Household income, highest educational attainment, and area-level deprivation (Townsend Deprivation Index).

**Main Outcomes and Measures:** Brain Age Gap (predicted minus chronological age, years) from T1-weighted (primary) and T2-FLAIR (secondary) MRI, derived with a deep learning model. Eleven risk factors and the Life’s Essential 8 cardiovascular health score were modeled as mediators; indirect effects were estimated in single-mediator and parallel models with 95% CIs from 5000 bootstrap resamples.

**Results:** Among 36 878 participants (mean [SD] age, 65.1 [7.7] years; 20 360 [55.2%] female), lower income and greater area deprivation were associated with a larger brain age gap: lowest vs highest income group, 0.35 years (95% CI, 0.22-0.47); most vs least deprived quartile, 0.33 years (95% CI, 0.23-0.43). Education showed weaker associations that differed in direction between imaging contrasts. Life’s Essential 8 score mediated 36% of the income (indirect effect, 0.031 [95% CI, 0.025-0.036]) and 17.9% of the area-deprivation (0.024 [95% CI, 0.019-0.029]) associations. In parallel models entering all risk factors, smoking was the largest mediator for both income (0.022 [95% CI, 0.016-0.028]) and area-deprivation (0.030 [95% CI, 0.023-0.037]). Alcohol intake offset the income association but contributed to the area-level deprivation association.

**Conclusions and Relevance:** Socioeconomic differences in brain age gap were partly accounted for by modifiable cardiovascular and lifestyle risk factors, with smoking being the single largest contributor. These findings identify modifiable cardiovascular risk factors as a substantial component of socioeconomic inequalities in brain aging.

**Key Points:** *Question:* Do modifiable risk factors statistically mediate socioeconomic differences in the brain age gap, and do the contributing factors differ across socioeconomic indicators?

*Findings:* In this cohort study of 36 878 UK Biobank participants, lower household income and greater area-level deprivation were associated with an older-appearing brain, whereas educational attainment was not consistently associated. Modifiable risk factors, particularly smoking, mediated part of these associations, and the mediating factors differed by socioeconomic indicator.

*Meaning:* Modifiable risk factors are candidate targets for reducing socioeconomic inequalities in brain aging, where individual and area-level disadvantage are associated with divergent risk profiles.

## Introduction

Socioeconomic deprivation is associated with accelerated aging^1,2^ and poorer health, including higher risk of stroke^3^ and dementia^4^. In the brain, accelerated aging can be quantified using the brain age gap (BAG), the difference between predicted and chronological age, where a greater BAG is associated with dementia, cognitive decline, and mortality^5–8^.

Although lower socioeconomic status (SES) has been associated with poorer brain health^9,10,11^, and, in smaller samples, with greater BAG^12^, this literature has typically operationalized SES using a single indicator, as a composite score, or as a covariate^10,13,14^. Yet, different SES indicators (household income, education, and area-level deprivation), may reflect distinct underlying mechanisms^15,16^, and treating them as interchangeable could obscure indicator-specific targets for intervention. Education and income, for example, show separable associations with cardiovascular health^17,18^ and differential associations with brain structure and cognition^19,20^. Likewise, individual and area-level deprivation capture personal and contextual disadvantage, respectively, and show independent associations with mortality^21^. Whether different SES indicators produce convergent or divergent associations with BAG has not been systematically compared.

Identifying which modifiable factors account for the association between SES and brain aging is relevant for prevention. Socioeconomic position is not readily modifiable at the individual level, whereas the lifestyle, cardiovascular (CV), and social-emotional factors through which it may operate are established intervention targets. An intermediate role for these factors is mechanistically plausible: deprivation has been linked to worse health outcomes through stress, allostatic load, and a patterned exposure to modifiable risk factors^22–24^. These same factors account for a substantial share of dementia risk across the life course^25^, and because they are both modifiable and socioeconomically patterned, they represent plausible contributors to inequalities in brain health.

Prior work has addressed parts of this pathway separately. Modifiable lifestyle and CV risk factors explain considerable variance in BAG, though without accounting for SES or conducting a formal mediation analysis^14^. Life’s Essential 8 (LE8), a validated composite of eight modifiable CV behaviors and biomarkers^26^ is associated with BAG, though with SES treated as a confounder^27^. LE8 also partially mediates the association between low SES and increased dementia incidence, but without a neuroimaging measure of brain aging^28^. Similar risk factors mediate the association between SES and structural brain volumes, though using a composite SES score and without investigation of BAG^13^. Social-emotional factors, such as infrequent social contact, fall outside LE8, yet are socioeconomically patterned and have been associated with higher brain age^29^. Taken together, these works establish a plausible basis for modifiable risk factors to mediate the relationship between low SES and accelerated brain aging.

Here we test whether LE8 mediates associations between three SES indicators (education, income, area-level deprivation) and BAG in UK Biobank participants in mid to late life (Figure 1). We then estimate the contribution of 11 individual lifestyle, CV, and social-emotional risk factors, several outside LE8, in parallel models that adjust for all other mediators. Together, these analyses quantify how much of each SES gradient in BAG is attributable to modifiable risk factors. Analysis code and documentation are available on the GitHub repository (https://github.com/rockNroll87q/SocioBrain).

**Figure 1.**
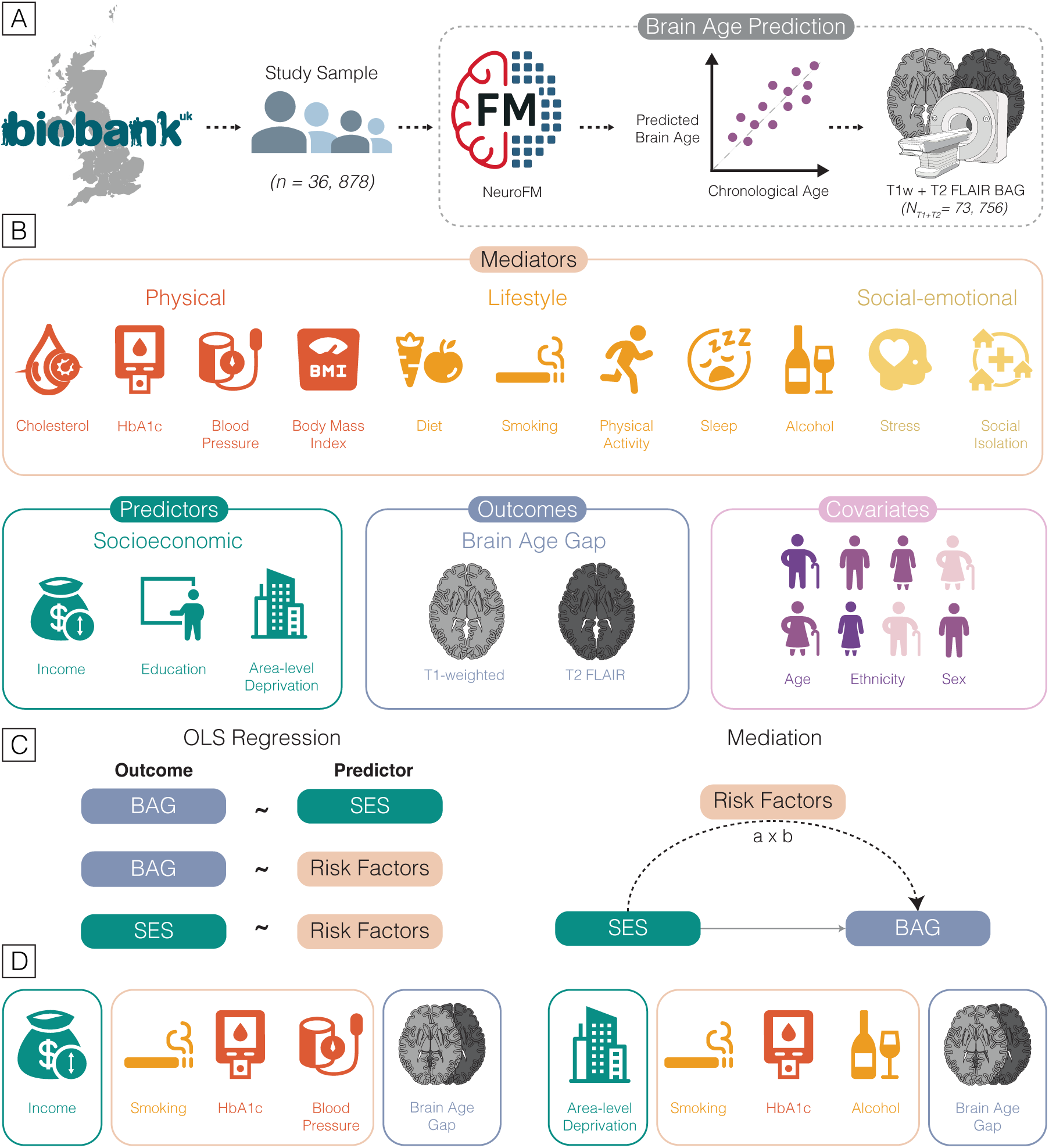
Study Overview. We tested whether modifiable cardiovascular and lifestyle risk factors mediate the association between socioeconomic status (SES) and brain age gap (BAG) in UK Biobank participants with structural brain MRI (n=36,878). (**A**) **Sample**. Brain age was estimated from T1-and T2-weighted MRI using NeuroFM^30^, yielding BAG estimates (predicted minus chronological age). SES and risk factor variables were then extracted for the imaging subsample. (**B) Variables.** SES exposures included household income, educational attainment, and area-level deprivation (Townsend Deprivation Index; TDI); risk factor variables included physical (cholesterol, HbA1c (blood glucose), body mass index (BMI), blood pressure), lifestyle (smoking, alcohol, sleep, diet, physical activity) and social-emotional (stress, social visit frequency) domains, together with the Life’s Essential 8 (LE8) composite score. **(C) Analysis.** Each exposure-outcome pair within each combination (SES → risk factor, SES → BAG, risk factor → BAG) was tested using an independent OLS regression. Mediation was assessed using the Baron & Kenny framework with SES as the exposure (X), risk factors as the mediator(s) (M), and BAG as the outcome (Y) in both single mediator (LE8) and parallel mediator (individual risk factors) models. **(D) Key parallel mediation results.** For the mediation of the income-BAG association, smoking, HbA1c, and blood pressure had the greatest positive indirect effects. For the mediation of the TDI-BAG association, smoking, HbA1c, and alcohol had the greatest positive indirect effects.

## Methods

### Data

#### UK Biobank

The UK Biobank (UKB) design and protocol have been described in detail elsewhere in open-access protocols^31^. Briefly, UKB recruited approximately 500,000 adults, aged 40 to 70 years at recruitment, from 2006 to 2010 across 22 UK assessment centers, with data collected through questionnaires, interviews, and anthropometric and biomedical measurements.

Ethical approval was obtained from the North West Multi-Centre Research Ethics Committee (reference number 11/NW/0382), and all participants provided written informed consent at baseline; the present analysis was conducted under UKB application 17689. The imaging substudy was conducted across four dedicated centers on Siemens Skyra 3T scanners^32^. Here, the analytic sample was restricted to participants with imaging data at first imaging visit.

#### SES

Three SES indicators were used as separate exposures, each recorded at baseline (mean [SD] interval to imaging, 9.31 [2.02] years): household income, highest educational qualification, and Townsend Deprivation Index (TDI)^33^, an area-based measure of material deprivation. In regression models, each indicator was entered categorically, income and education as collapsed categories and TDI as quartiles, with the most advantaged group (highest income, degree qualification, least deprived quartile, respectively) set as the reference. In mediation models, each indicator was entered continuously (income as a five-level ordinal, education as a 7-level ordinal, and TDI z-scored within-sample). Category totals, definitions, and mappings are provided in eMethods 1 and eFigure 1. In sensitivity analyses, the Index of Multiple Deprivation (England only) replaced TDI, which was handled identically.

### Risk factors

#### Individual physical, lifestyle, and social-emotional risk factors

Risk factors were assessed at baseline visit. Smoking (pack-years), alcohol (units/week), BMI, glycated hemoglobin (HbA1c), and systolic and diastolic blood pressure were z-scored. Pack-years smoking was log-transformed before standardization owing to skew. Unhealthy diet was scored on a 9-point scale and physical inactivity was derived by reverse coding total activity, oriented so that higher values represent worse health. Suboptimal sleep, high cholesterol (non-high-density lipoprotein; non-HDL), infrequent social visits, and stress were binarized. Cholesterol-and blood pressure-lowering medication use was assessed at both baseline and imaging and incorporated into the relevant biomarker definitions. Full coding details are provided in eMethods 2, Risk Factor Coding).

#### Composite CV health - Life’s Essential 8

LE8 was calculated as per Petermann-Rocha et al^34^, where it had been adapted for the UKB. The LE8 score includes eight components (smoking status, BMI, physical activity, diet, non-HDL cholesterol, blood pressure, HbA1c, and sleep duration), each scored from 0-100. Participants with any missing components were treated as missing for the total score, which was then z-scored within the imaging subsample.

#### Brain Age Gap estimation

Brain age was estimated using NeuroFM, a 3D convolutional neural network pretrained on 100,000 synthetic T1-weighted brain MRI volumes to predict chronological age and morphometry^30^. Fine-tuning to generate separate T1-weighted (MPRAGE; hereafter T1) and T2-weighted fluid-attenuated inversion recovery (FLAIR; hereafter T2) brain age estimates was performed in previous work using an independent model development sample of 7,700 UK Biobank participants (15,400 volumes) selected on health criteria^30^. Fine-tuning participants were excluded from downstream analyses, yielding an analytic sample of 36,878 participants with held-out estimates. BAG was defined per modality as predicted minus chronological age at imaging (years), with positive values indicating an older appearing brain (model performance, T1: MAE 3.38 years, *r* = 0.856; T2: MAE 4.39 years; *r* = 0.764, eMethods 3, eFigure 2 in Supplement 1).

## Statistical Analysis

Analyses were performed in Python v3.12.3 (statsmodels, pandas, numpy) and followed STROBE and AGReMA reporting guidelines. All models were adjusted for age, sex, and ethnicity. Age at baseline and at imaging were z-scored and treated as continuous. Ethnicity was dichotomized as white vs non-white given UKB’s predominantly white sample. Tests were two-sided; significance was defined as Benjamini-Hochberg FDR-adjusted *P* < 0.05. The primary outcome was T1 BAG, with T2 BAG as a secondary outcome. Sensitivity analyses are described in eMethods 4.

### Association Analyses

Three families of association were estimated: SES with BAG, risk factors with BAG, and SES with risk factors. Each exposure-outcome pair was modeled with separate ordinary least squares regression and fit on complete cases, so sample size varied across models. FDR correction was applied within each family.

### Mediation analysis

Mediation models followed the framework of Baron and Kenny^35^, with bootstrapped indirect effects. For each exposure-mediator-outcome combination, three regression models estimated the total effect (*c*), paths *a* and *b*, and the direct effect (*c’).* The indirect effect (*a x b*) was tested with 95% confidence intervals from 5,000 percentile bootstrap resamples. The proportion mediated (*ab/c)* is reported for single-mediator models only (eMethods 4). Single-mediator models estimated the contribution of overall CV health (LE8); parallel models entered all risk factors simultaneously to estimate each factor’s independent contribution.

Estimates from the two are not directly comparable. FDR was applied across all indirect-effect tests at a = 0.05. Given the cross-sectional exposure-mediator relationship, estimates reflect statistical association under assumptions of no unmeasured confounding rather than causal effects.

## Results

### Demographics

The sample comprised 36,878 participants (mean [SD] age, 65.1 [7.7] years, 55.2% female; Table 1). All models included age, sex, and ethnicity.

**Table 1.** Characteristics of 36 878 UK Biobank Participants with BAG predictions ^a^.

| Variable | Value | Missing |
| --- | --- | --- |
| N | 36 878 |  |
| **Demographics** |  |  |
| Age at first imaging visit (years) (SD) | 65.1 (7.7) | 0 (0.0%) |
| Female (%) | 20 360 (55.2%) | 0 (0.0%) |
| White ethnicity (%) <sup>b</sup> | 35 715 (97.1%) | 106 (0.3%) |
| **Brain Age** |  |  |
| Brain age gap, T1 (years) (SD) | -1.0 (4.0) | 0 (0.0%) |
| Brain age gap, T2 (years) (SD) | -1.7 (5.1) | 0 (0.0%) |
| **Socioeconomic Status** |  |  |
| Education (%) |  | 613 (1.7%) |
| None | 2596 (7.2%) |  |
| Secondary | 7224 (19.9%) |  |
| Post-secondary | 10 297 (28.4%) |  |
| Degree-level | 16 148 (44.5%) |  |
| Household income (£) (%) |  | 3675 (10.0%) |
| < 18 000 | 3994 (12.0%) |  |
| 18 000–30 999 | 7507 (22.6%) |  |
| 31 000–51 999 | 10 015 (30.2%) |  |
| 52 000–100 000 | 9153 (27.6%) |  |
| > 100 000 | 2534 (7.6%) |  |
| Townsend Deprivation Index (SD)[Range] | -1.83 (2.74) [-6.26, 10.10] | 31 (0.1%) |
| **Risk factors** |  |  |
| LE8 score, 0-100 (SD)<sup>c</sup> | 67.9 (10.9) | 9572 (26.0%) |
| Smoking status (%) |  | 93 (0.3%) |
| Never | 20 453 (55.6%) |  |
| Former | 13 721 (37.3%) |  |
| Current | 2611 (7.1%) |  |
| Pack-years, ever-smokers (n=10,998) [IQR]<sup>d</sup> | 15.0 [7.8, 25.5] | 5334/16 332 (32.7%) |
| Alcohol (units/week) [IQR] | 15.3 [8.5, 26.7] | 6 848 (18.6%) |
| Suboptimal sleep (<7 or >9 h) (%) | 8443 (23.0%) | 99 (0.3%) |
| Physical activity (MET min/week) [IQR] | 1533 [678, 3040] | 1 048 (2.8%) |
| Healthy diet score (0-9) (SD) | 4.5 (1.4) | 12 (<0.1%) |
| Body mass index (kg/m<sup>2</sup>) (SD) | 27.2 (4.3) | 56 (0.2%) |
| Non-HDL cholesterol (mg/dL) (SD) | 165.9 (39.9) | 5485 (14.9%) |

**Table 1. Continued**
| Variable | Value | Missing |
| --- | --- | --- |
| Systolic blood pressure (mmHg) (SD) | 135.6 (17.7) | 2265 (6.1%) |
| Diastolic blood pressure (mmHg) (SD) | 81.8 (9.9) | 2265 (6.1%) |
| HbA1c (%) (SD) | 5.4 (0.5) | 2712 (7.4%) |
| Frequently tense/restless (yes, %) | 8873 (24.7%) | 896 (2.4%) |
| Infrequent friend/family visits (yes, %) | 2712 (7.5%) | 601 (1.6%) |

Abbreviations: BAG, brain age gap; BMI, body mass index; HbA1c, glycated hemoglobin; IQR, interquartile range; LE8, Life's Essential 8, MET, metabolic equivalent of task; SD, standard deviation; TDI, Townsend Deprivation Index. SI conversion factors: non-HDL cholesterol to mmol/L, multiply by 0.0259; HbA1c to proportion of total hemoglobin, multiply by 0.01.
<sup>a</sup> Data are presented as mean (SD), median [IQR], or No. (%) unless otherwise indicated. Percentages are calculated among participants with non=missing data.
<sup>b</sup> Race and ethnicity were self-reported by participants at the baseline assessment from fixed categories defined by UK Biobank.
<sup>c</sup> LE8 is scored from 0 to 100. Missingness is greater than other risk factors as LE8 requires presence of all components.
<sup>d</sup> Pack-years were assessed only among ever smokers (n = 16 332). The reported median is based on the 10 998 ever smokers with non-missing data, and missingness is expressed as a proportion of ever smokers.

### Socioeconomic status and Brain Age Gap

In individual regression models, T1 and T2 estimates agreed in sign for 19 of 22 exposures; all exceptions were education contrasts (eFigure 3 in Supplement 1). T1 estimates are reported below except where estimates diverged in sign or statistical significance.

Lower income was associated with greater BAG (ie, older appearing brain) in a graded pattern, where the lowest income group (<£18,000) showed the largest difference (b = 0.348 years, 95% CI, 0.223 to 0.472). The £31-52,000 group did not differ from the > £52,000 reference (Figure 2a, eTable 1).

**Figure 2.**
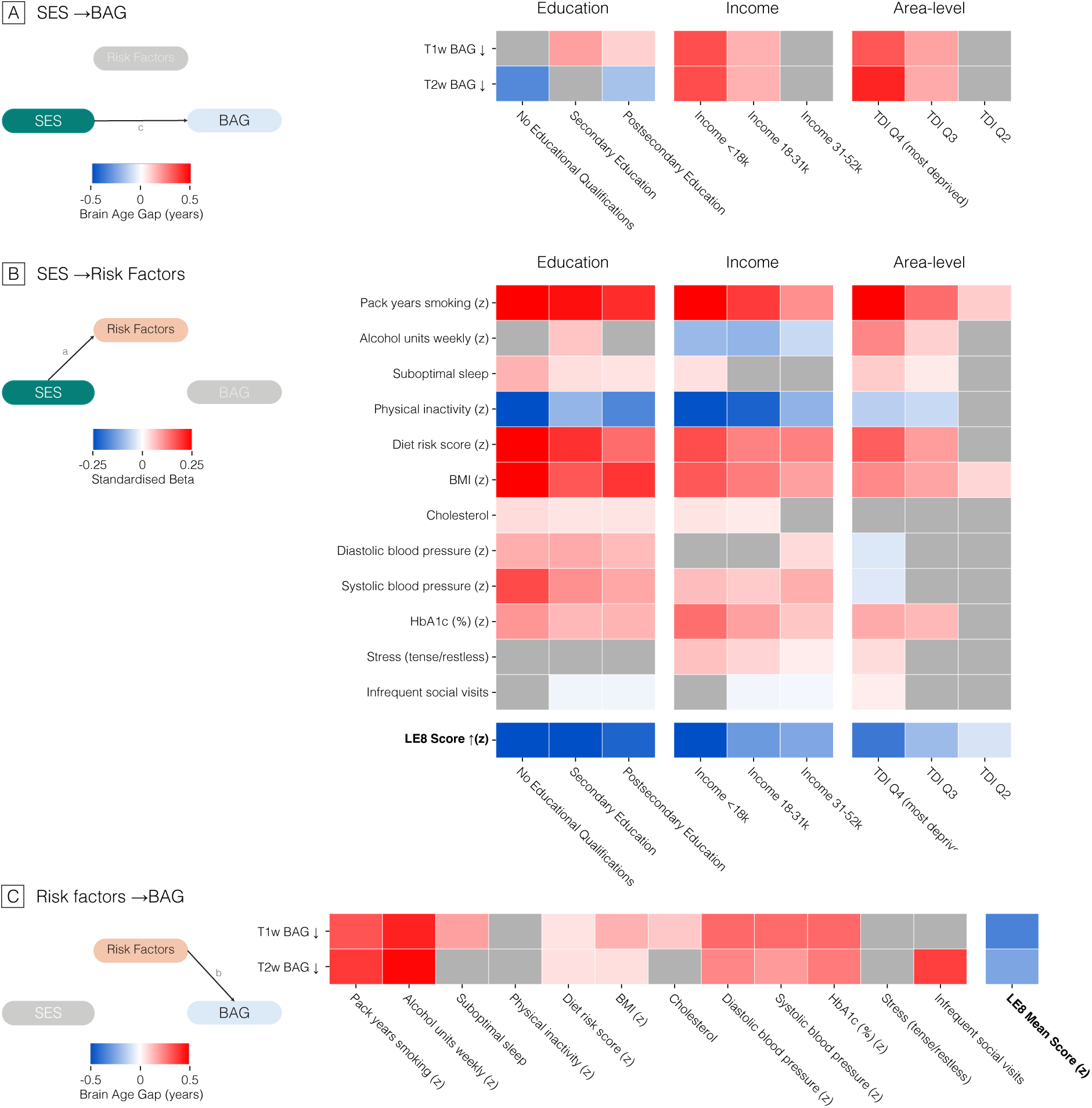
Socioeconomic status is associated with brain age gap (A) and with cardiovascular and lifestyle risk factors (B), which are in turn associated with brain age gap (C). Throughout, each cell is one independent OLS model of a single outcome (rows) on a single exposure (columns), adjusted for age, sex, and ethnicity. Categorical SES indicators (education levels, income brackets, TDI quartiles) are dummy coded contrasts against the high-SES reference category (university degree; income > £52,000; TDI Q1). Risk factors are ordered by domain: lifestyle (smoking, alcohol, sleep, physical activity, diet), physical (BMI, cholesterol, blood pressure, HbA1c), social-emotional (stress, social visit frequency), plus the LE8 composite score; all are z scored except cholesterol, stress, and infrequent social visits, which are binarized. Cell color shows the β coefficient: red is a higher value of the outcome measure, and blue a lower value. Arrows beside row labels mark the healthier direction, so red indicates the more adverse direction for every outcome except LE8, for which higher scores are healthier. Color scales differ between panels; see color bars. Grey cells are non-significant after Benjamini-Hochberg FDR correction applied across all tests within each panel. (**A**) BAG on each SES indicator. Coefficients are the difference in BAG (years) for each SES category relative to the reference category (n = 15,647-26,372). (**B**) Each risk factor on each SES indicator. Coefficients are standardized (SD units of the risk factor) for each SES category relative to the reference (n = 11,841-26,372). **(C)** BAG on each risk factor. Coefficients are the difference in BAG (years) per 1-SD higher exposure, or for the adverse healthy category in binarized exposures (n = 27,259-36,772). BAG, brain age gap; BMI, body mass index; FDR, false discovery rate; LE8, Life’s Essential 8; OLS, ordinary least squares; SD, standard deviation; SES, socioeconomic status; TDI, Townsend Deprivation Index.

The same graded pattern was observed for TDI, with the largest difference between the most (Q4) and least (Q1) deprived quartiles (b = 0.329 years, 95% CI, 0.232 to 0.426) (Figure 2a). Results were consistent using IMD (eFigure 4).

Education estimates diverged by modality. Compared with university education, secondary (b = 0.184 years, 95% CI, 0.092 to 0.276) and postsecondary education (b = 0.094 years, 95% CI, 0.013 to 0.175) were associated with greater T1 BAG. For T2 BAG, no qualifications (b =-0.333 years, 95% CI,-0.519 to-0.148) and postsecondary education (b =-0.173 years, 95% CI,-0.281 to-0.066) were associated with lower BAG. The education association differed by sex for T2 BAG (*F*₃,₃₆₁₆₃ = 4.45; *P* =.004.) but not by age band (P =.40); T1 estimates were directionally similar but nonsignificant (P =.11) (eFigure 5, eTable 2 in Supplement 1).

### Socioeconomic status and risk factors

Lower SES was associated with lower LE8 scores (worse cardiovascular health) across all three indicators (No educational qualifications, β =-0.437, 95% CI,-0.483 to-0.391; < £18k, β =-0.264, 95% CI,-0.305 to-0.223; TDI Q4, β =-0.190, 95% CI,-0.222 to-0.159) (Figure 2b). SES indicators showed a graded association with LE8, where the lowest SES groups were associated with the lowest LE8 scores (eTable 3 in Supplement 1).

Lower SES was associated with higher smoking, BMI, blood pressure, HbA1c, worse diet, and worse sleep, except for the most deprived quartile (Q4), which was associated with lower blood pressure. Results were consistent using IMD, except for blood pressure (eFigure 6).

Lower income was associated with lower alcohol intake (income < 18k, b =-0.098, 95% CI, - 0.143 to-0.054), whereas greater area-level deprivation was associated with higher intake (TDI Q4, b = 0.119, 95% CI, 0.086 to 0.152). Lower SES was associated with greater total physical activity. Lower education and income, but not area-level deprivation, were associated with higher non-HDL cholesterol. Lower income and greater area-level deprivation were weakly associated with higher stress, with mixed results for education.

### Risk Factors and Brain Age Gap

Higher LE8 score was associated with lower BAG (b =-0.354 years per SD, 95% CI,-0.395 to-0.312) (Figure 2c). Higher smoking, alcohol consumption, blood pressure, HbA1c, BMI, and worse diet were associated with greater BAG across both modalities (eTable 4 in Supplement 1). Infrequent social contact was associated with greater T2, but not T1, BAG, and higher cholesterol and suboptimal sleep with higher T1, but not T2, BAG. Neither stress nor physical inactivity was associated with BAG.

### Single-Mediator Model (Life’s Essential 8)

Throughout, the sign of income estimates was reversed for presentation so that positive values, as for TDI, denote greater socioeconomic disadvantage. Indirect effects (a ‘b) are reported with 95% CIs. In single mediator models, LE8 partially mediated both the income-BAG (0.031 [95% CI, 0.025 to 0.036]) and TDI-BAG (0.024 [95% CI, 0.019 to 0.029]) associations (Figure 3). Indirect effects were larger for income than TDI, and for T1 than T2. The same T1/T2 pattern was observed for IMD (eFigure 7, eTable 5 in Supplement 1).

**Figure 3.**
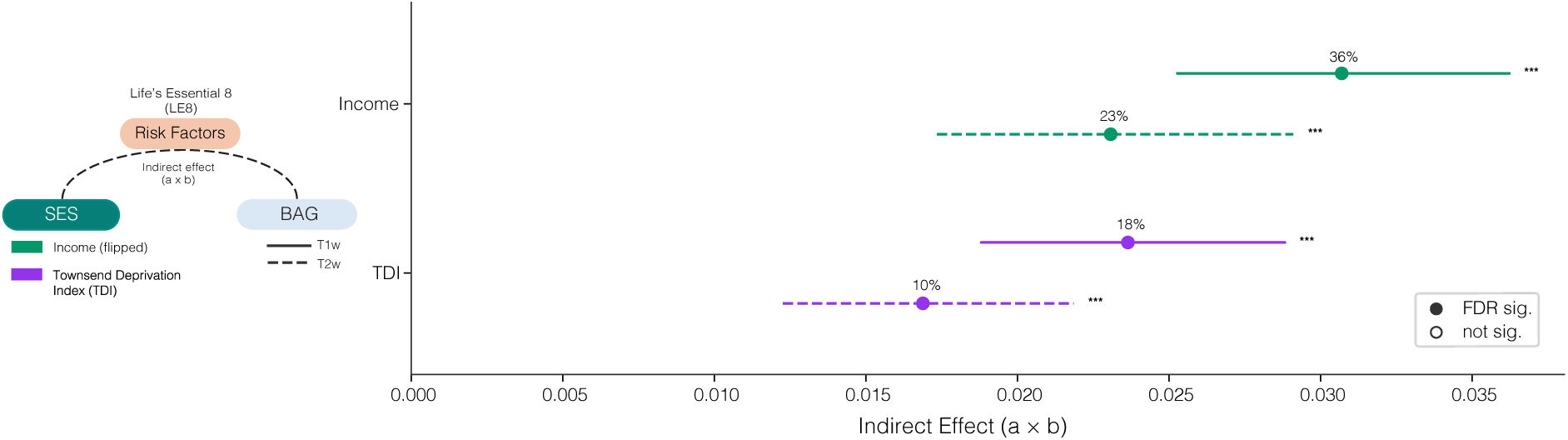
LE8 partially mediates the SES-BAG association. Forest plot of indirect effects (a x b) from the single mediator models testing the LE8 composite score as a mediator of the association between SES (income and TDI) and BAG. Each row is one X → M → Y pathway; points are point estimates and horizontal bars are 5,000-iteration percentile bootstrap 95% confidence intervals. Pathways are colored by SES exposure (purple, TDI; green, Income, sign-flipped). Indirect effects for income were sign flipped so that, for both SES indicators, a positive indirect effect relates to lower SES being associated with higher BAG via worse CV health. Filled markers indicate pathways significant after Benjamini-Hochberg FDR correction (applied across all X∼M∼Y combinations in the run); open markers are non-significant. *** *q* < 0.001, ** *q* < 0.01, * *q* < 0.05. Per-pathway proportion mediated is annotated above each bar. Models adjusted for age, sex, and ethnicity (n = 27,199). BAG, brain age gap; CI confidence interval; FDR, false discovery rate; LE8, Life’s Essential 8; SES, socioeconomic status.

### Parallel Mediation Models

In the parallel models, where all risk factors were entered simultaneously, smoking showed the largest indirect effect for income-BAG (0.022 [95% CI, 0.016 to 0.028]) (Figure 4a), followed by HbA1c (T1: 0.009 [95% CI, 0.006 to 0.013]), systolic blood pressure (T1: 0.009 [95% CI, 0.006 to 0.013]) and BMI (T1 only). Other lifestyle (sleep, diet), and social-emotional risk factors were nonsignificant (eTable 6 and eTable 7).

**Figure 4.**
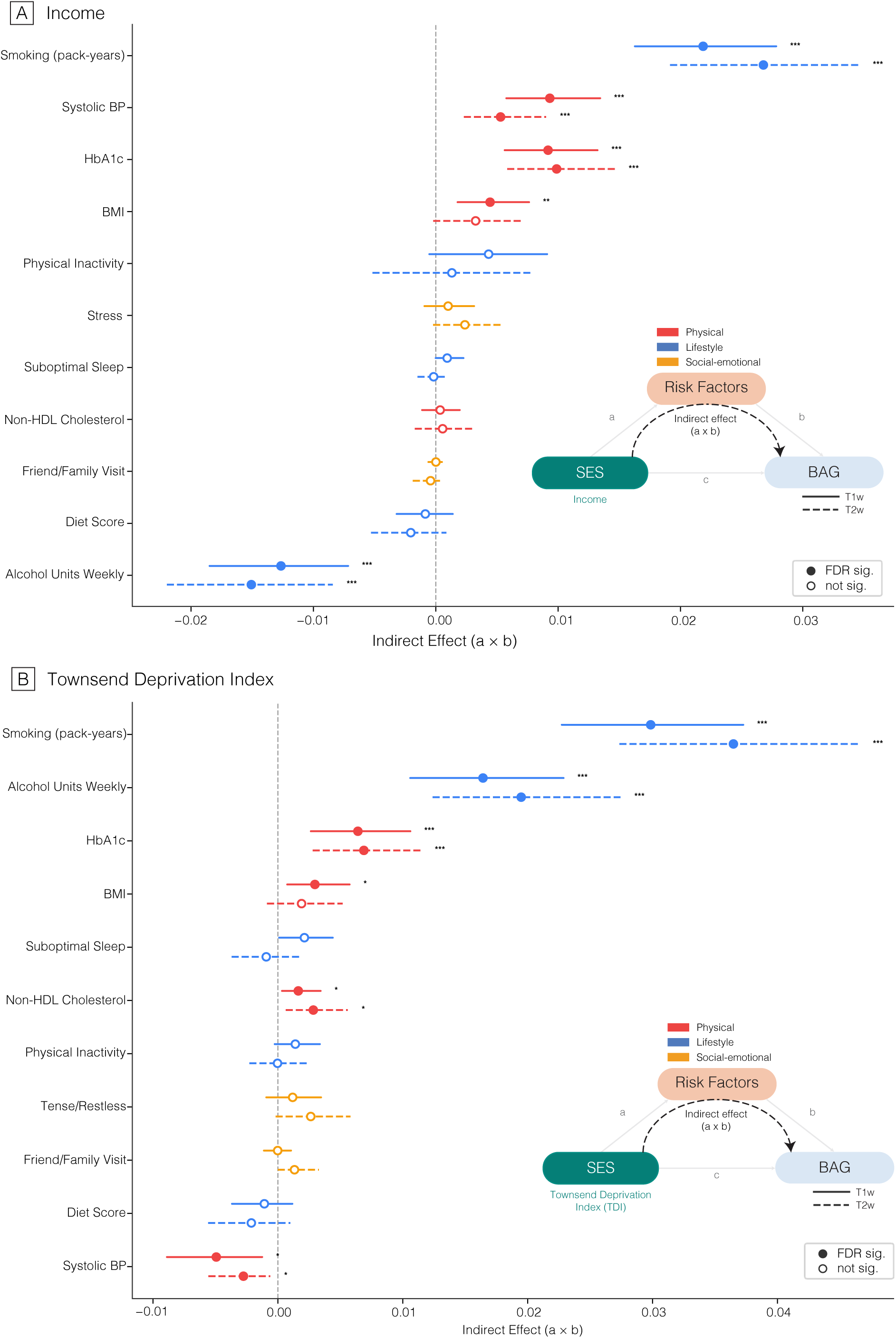
Smoking carries most of the income → BAG association (A), while smoking and alcohol carry most of the TDI → BAG association (B). Forest plot of specific indirect effects (*a_k_ x b_k_)* from the parallel mediation model of SES → BAG, with all eleven risk factors entered simultaneously as mediators. Panels show (a) income → BAG and (b) TDI → BAG. Within each panel, solid lines show T1 BAG and dashed lines show T2 BAG. Each row is one mediator; points are point estimates and horizontal bars are 5000-iteration percentile bootstrap 95% confidence intervals. Mediators are colored by domain (red, physical; blue, lifestyle; yellow, social-emotional) and ordered within each panel by signed indirect effect (row order differs between panels. Filled markers indicate mediation paths significant after Benjamini-Hochberg FDR correction (applied across all X∼M∼Y combinations in the parallel mediation run, eg, income-T1); open markers are non-significant. \*\*\**q* < 0.001, \*\**q* < 0.01, \**q* < 0.05. Per mediator proportion mediated is annotated to the right of each bar. Models adjusted for age, sex, and ethnicity. **(A)** Income → BAG (n = 16,886). Income was sign-flipped so that a positive specific indirect effect corresponds to lower income → higher BAG via that mediator; the negative indirect effect for alcohol therefore reflects the positive income–alcohol association in this cohort. **(B)** TDI → BAG (n = 18,244). Higher TDI indicates greater area-level deprivation, so a positive specific indirect effect corresponds to greater deprivation being associated with higher BAG via that mediator. BAG, brain age gap; BMI, body mass index; BP, blood pressure; CI, confidence interval; FDR, false discovery rate; SES, socioeconomic status; TDI, Townsend Deprivation Index.

Alcohol showed positive indirect effects for TDI (0.016 [95% CI, 0.011 to 0.023]) (Figure 4b), but negative indirect effects for income (-0.013 [95% CI,-0.018 to-0.007]; suppression effect; Figure 4a). The counterintuitive mediation of income-BAG reflects income’s positive association with alcohol intake (higher income, higher alcohol intake) (Figure 2b). Here, alcohol intake counteracts part of the lower BAG otherwise associated with higher income. Negative indirect effects were also observed for blood pressure’s mediation of TDI-BAG, this time reflecting TDI’s negative association with blood pressure (higher area deprivation, lower blood pressure) (Figure 2b). Like income, HbA1c showed the largest indirect effect among physical risk factors for TDI-BAG, followed by BMI (T1 only) and cholesterol. Other lifestyle (sleep, diet), and social emotional risk factors showed near zero indirect effects (eTable8 and eTable9). Education parallel models are shown in eFigure 8.

## Discussion

In this population-based study, lower income and greater area-level deprivation were associated with an older appearing brain, with a substantial share of each gradient accounted for by modifiable risk factors. Smoking was the largest single mediator for every SES indicator across both imaging contrasts, followed by HbA1c and blood pressure. Alcohol intake acted in opposite directions across indicators, suppressing the income association while contributing to the deprivation association; a divergence that indicates individual and area-level measures of disadvantage cannot be treated as interchangeable. Overall CV health, summarized by LE8, mediated 36% of the income association and 17.9% of the area-deprivation association with BAG. To our knowledge, this is the first study to decompose the association between SES and BAG into modifiable risk factor contributions across multiple SES indicators.

Previous studies have reported associations between income and brain structure^19^, and income and BAG^36^. As in our findings, associations between education and BAG are more varied^14,36^, and the broader evidence for education and age related decline and structural change is similarly mixed^37,38^. Our finding that lower educational attainment was associated with lower T2 BAG should therefore be interpreted with caution. The association reversed in direction between contrasts in the same participants and was present only in females (interaction *P* =.004). This pattern is more consistent with differential selection (participation in the imaging substudy is itself socioeconomically patterned^39^) or contrast-specific measurement, than with a protective effect of lower education.

The LE8 score was associated with BAG across both imaging contrasts. Within LE8, several components emerged as major contributors including smoking, blood pressure, HbA1c, and BMI. This is in line with the existing midlife vascular risk and brain aging literature^36,40,41^. The null association between physical activity and BAG may reflect the known U-shaped activity-BAG association^42^. Lower SES was also associated with greater physical activity.

Both observations may reflect the physical activity paradox, whereby lower SES individuals accumulate the majority of their physical activity through occupational means rather than leisure^43^, and occupational activity, being sustained, lower in intensity, and accompanied by insufficient recovery, does not confer the same cardiometabolic benefits^43,44^.

LE8 partially mediated SES-BAG associations, most strongly for income-T1 BAG (36%), comparable to its mediation of the SES-dementia association^28^. However, these results show that, particularly for TDI-T2 (9.9%), a large portion of the association runs through pathways that LE8 does not capture.

Across the parallel mediation models, smoking was consistently the largest single mediator, for each SES indicator and imaging contrast. This reflects known associations between smoking and worse brain health^45^ and BAG^41^, highlighting smoking as a leading candidate factor through which socioeconomic disadvantage relates to accelerated brain aging. Because smoking is modifiable, strongly socioeconomically patterned, and a considerable risk factor for dementia^25^, it is a plausible priority for research aimed at narrowing socioeconomic inequalities in brain health.

Higher alcohol intake was associated with higher income, consistent with previous UKB literature^46,47^, and with higher BAG, reflecting its adverse effects on brain health and structure^48^. This produces opposing indirect effects across indicators. In the income model alcohol is a negative, suppressing mediator, because higher income (higher SES) is associated with higher intake and higher intake with higher BAG. In the TDI model it is a positive mediator, as greater area deprivation (lower SES) is associated with lower intake.

This study has several limitations. First, SES and risk factors were assessed concurrently, so the assumed ordering of exposure and mediator could not be verified. BAG was measured on a single occasion (mean 9.31[2.02] years after baseline), and no baseline measure of brain health was available, so reverse causation cannot be excluded. The results therefore describe an associational decomposition rather than causal effects, and no causal claims are made.

Second, the UKB imaging subsample carries a well-documented healthy-volunteer bias, with participants being healthier, wealthier, and less ethnically diverse than the general UK population^39,49^. Third, indirect effects depend on the reliability with which mediators are measured. The social-emotional factors were single-item self-reports, whereas LE8 summarizes eight measured components, so their null indirect effects may reflect measurement error rather than absence of mediation. Both selection into the imaging cohort and imperfect mediator measurement attenuate estimates, so indirect effects are likely lower bounds. Finally, BAG is a global, whole-brain summary measure. While it is a validated marker of overall brain health^6–8^, it cannot localize which regions drive the observed associations. Region-specific measures, typically derived from segmentation and surface reconstruction pipelines^50–53^, may offer greater anatomical specificity and represent a useful complement in future work.

In this cohort, socioeconomic gradients in BAG were partially mediated by modifiable CV and lifestyle risk factors, with smoking the largest single contributor across all indicators and both contrasts. These modifiable factors represent candidate priorities for research aimed at reducing socioeconomic inequalities in brain health. We also found that separate SES indicators differ in their associations with both risk factors and BAG, suggesting that SES indicators should not be treated as interchangeable in future epidemiological work.

## Data Availability

The data used in this study were obtained from UK Biobank under application number 17689. UK Biobank data are available to bona fide researchers through an application process managed by UK Biobank. The authors are not permitted to redistribute individual-level data. Analysis code supporting this study is available at https://github.com/rockNroll87q/SocioBrain

## References

1. Steptoe A, Zaninotto P. Lower socioeconomic status and the acceleration of aging: An outcome-wide analysis. Proc Natl Acad Sci. 2020;117(26):14911–14917. doi:10.1073/pnas.1915741117

2. Lawrence KG, Kresovich JK, O’Brien KM, et al. Association of Neighborhood Deprivation With Epigenetic Aging Using 4 Clock Metrics. JAMA Netw Open. 2020;3(11):e2024329. doi:10.1001/jamanetworkopen.2020.24329

3. Marshall IJ, Wang Y, Crichton S, McKevitt C, Rudd AG, Wolfe CDA. The effects of socioeconomic status on stroke risk and outcomes. Lancet Neurol. 2015;14(12):1206–1218. doi:10.1016/S1474-4422(15)00200-8

4. Cadar D, Lassale C, Davies H, Llewellyn DJ, Batty GD, Steptoe A. Individual and Area-Based Socioeconomic Factors Associated With Dementia Incidence in England: Evidence From a 12-Year Follow-up in the English Longitudinal Study of Ageing. JAMA Psychiatry. 2018;75(7):723–732. doi:10.1001/jamapsychiatry.2018.1012

5. Gaser C, Franke K, Klöppel S, Koutsouleris N, Sauer H. BrainAGE in Mild Cognitive Impaired Patients: Predicting the Conversion to Alzheimer’s Disease. PLoS ONE. 2013;8(6):e67346. doi:10.1371/journal.pone.0067346

6. Cole JH, Ritchie SJ, Bastin ME, et al. Brain age predicts mortality. Mol Psychiatry. 2018;23(5):1385–1392. doi:10.1038/mp.2017.62

7. Zhang R, Yi F, Mao H, Huang Z, Wang K, Zhang J. Brain age gap as a predictive biomarker that links aging, lifestyle, and neuropsychiatric health. Commun Med. 2025;5(1):441. doi:10.1038/s43856-025-01100-5

8. Elliott ML, Belsky DW, Knodt AR, et al. Brain-age in midlife is associated with accelerated biological aging and cognitive decline in a longitudinal birth cohort. Mol Psychiatry. 2021;26(8):3829–3838. doi:10.1038/s41380-019-0626-7

9. Hunt JFV, Vogt NM, Jonaitis EM, et al. Association of Neighborhood Context, Cognitive Decline, and Cortical Change in an Unimpaired Cohort. Neurology. 2021;96(20):e2500–e2512. doi:10.1212/WNL.0000000000011918

10. Hunt JFV, Buckingham W, Kim AJ, et al. Association of Neighborhood-Level Disadvantage With Cerebral and Hippocampal Volume. JAMA Neurol. 2020;77(4):451. doi:10.1001/jamaneurol.2019.4501

11. Krueger KR, Desai P, Beck T, et al. Lifetime Socioeconomic Status, Cognitive Decline, and Brain Characteristics. JAMA Netw Open. 2025;8(2):e2461208. doi:10.1001/jamanetworkopen.2024.61208

12. Busby N, Newman-Norlund S, Sayers S, et al. Lower socioeconomic status is associated with premature brain aging. Neurobiol Aging. 2023;130:135–140. doi:10.1016/j.neurobiolaging.2023.06.012

13. Beydoun MA, Beydoun HA, Fanelli-Kuczmarski MT, et al. Uncovering mediational pathways behind racial and socioeconomic disparities in brain volumes: insights from the UK Biobank study. GeroScience. 2025;47(2):1837–1858. doi:10.1007/s11357-024-01371-1

14. Stoitsas K, Bakx P, Voortman T, Yu J, Roshchupkin G, Bos D. Contributions of lifestyle, education, and cardiovascular risk factors to the brain age gap. Aging Brain. 2025;8:100149. doi:10.1016/j.nbas.2025.100149

15. Braveman PA, Cubbin C, Egerter S, et al. Socioeconomic Status in Health ResearchOne Size Does Not Fit All. JAMA. 2005;294(22):2879–2888. doi:10.1001/jama.294.22.2879

16. Geyer S, Hemström Ö, Peter R, Vågerö D. Education, income, and occupational class cannot be used interchangeably in social epidemiology. Empirical evidence against a common practice. J Epidemiol Community Health. 2006;60(9):804–810. doi:10.1136/jech.2005.041319

17. Duan MJF, Zhu Y, Dekker LH, et al. Effects of Education and Income on Incident Type 2 Diabetes and Cardiovascular Diseases: a Dutch Prospective Study. J Gen Intern Med. 2022;37(15):3907–3916. doi:10.1007/s11606-022-07548-8

18. Newman DB, Gordon AM, Mendes WB. Income and education show distinct links to health and happiness in daily life. Nat Hum Behav. 2025;9(11):2299–2312. doi:10.1038/s41562-025-02264-9

19. Lotze M, Domin M, Schmidt CO, Hosten N, Grabe HJ, Neumann N. Income is associated with hippocampal/amygdala and education with cingulate cortex grey matter volume. Sci Rep. 2020;10(1):18786. doi:10.1038/s41598-020-75809-9

20. Walhovd KB, Fjell AM, Wang Y, et al. Education and Income Show Heterogeneous Relationships to Lifespan Brain and Cognitive Differences Across European and US Cohorts. Cereb Cortex. 2022;32(4):839–854. doi:10.1093/cercor/bhab248

21. Ribeiro AI, Fraga S, Severo M, et al. Association of neighbourhood disadvantage and individual socioeconomic position with all-cause mortality: a longitudinal multicohort analysis. Lancet Public Health. 2022;7(5):e447–e457. doi:10.1016/S2468-2667(22)00036-6

22. Seeman T, Epel E, Gruenewald T, Karlamangla A, McEwen BS. Socio-economic differentials in peripheral biology: Cumulative allostatic load. Ann N Y Acad Sci. 2010;1186(1):223–239. doi:10.1111/j.1749-6632.2009.05341.x

23. Stringhini S, Sabia S, Shipley M, et al. Association of Socioeconomic Position With Health Behaviors and Mortality. JAMA. 2010;303(12):1159–1166. doi:10.1001/jama.2010.297

24. Businelle MS, Mills BA, Chartier KG, Kendzor DE, Reingle JM, Shuval K. Do stressful events account for the link between socioeconomic status and mental health? J Public Health. 2014;36(2):205–212. doi:10.1093/pubmed/fdt060

25. Livingston G, Huntley J, Liu KY, et al. Dementia prevention, intervention, and care: 2024 report of the Lancet standing Commission. The Lancet. 2024;404(10452):572–628. doi:10.1016/S0140-6736(24)01296-0

26. Lloyd-Jones DM, Allen NB, Anderson CAM, et al. Life’s Essential 8: Updating and Enhancing the American Heart Association’s Construct of Cardiovascular Health: A Presidential Advisory From the American Heart Association. Circulation. 2022;146(5):e18–e43. doi:10.1161/CIR.0000000000001078

27. Feng L, Ye Z, Pan Y, et al. Adherence to life’s essential 8 is associated with delayed white matter aging. eBioMedicine. 2025;115:105723. doi:10.1016/j.ebiom.2025.105723

28. Van Der Heide FCT, Valeri L, Dugravot A, et al. Role of cardiovascular health factors in mediating social inequalities in the incidence of dementia in the UK: two prospective, population-based cohort studies. eClinicalMedicine. 2024;70:102539. doi:10.1016/j.eclinm.2024.102539

29. de Lange AMG, Kaufmann T, Quintana DS, et al. Prominent health problems, socioeconomic deprivation, and higher brain age in lonely and isolated individuals: A population-based study. Behav Brain Res. 2021;414:113510. doi:10.1016/j.bbr.2021.113510

30. Dibble A, Dalby C, Sevegnani M, et al. NeuroFM: Toward Precision Neuroimaging with Foundation Models for Individualized Brain Health Estimation. medRxiv. Published online April 1, 2026:2026.03.27.26349489. doi:10.64898/2026.03.27.26349489

31. Sudlow C, Gallacher J, Allen N, et al. UK Biobank: An Open Access Resource for Identifying the Causes of a Wide Range of Complex Diseases of Middle and Old Age. PLOS Med. 2015;12(3):e1001779. doi:10.1371/journal.pmed.1001779

32. Alfaro-Almagro F, Jenkinson M, Bangerter NK, et al. Image processing and Quality Control for the first 10,000 brain imaging datasets from UK Biobank. NeuroImage. 2018;166:400–424. doi:10.1016/j.neuroimage.2017.10.034

33. Townsend P, Phillimore P, Beattie A. Health and Deprivation: Inequality and the North. Croom Helm; 1988.

34. Petermann-Rocha F, Deo S, Celis-Morales C, et al. An Opportunity for Prevention: Associations Between the Life’s Essential 8 Score and Cardiovascular Incidence Using Prospective Data from UK Biobank. Curr Probl Cardiol. 2023;48(4):101540. doi:10.1016/j.cpcardiol.2022.101540

35. Baron RM, Kenny DA. The moderator–mediator variable distinction in social psychological research: Conceptual, strategic, and statistical considerations. J Pers Soc Psychol. 1986;51(6):1173–1182. doi:10.1037/0022-3514.51.6.1173

36. Jawinski P, Markett S, Drewelies J, et al. Linking Brain Age Gap to Mental and Physical Health in the Berlin Aging Study II. Front Aging Neurosci. 2022;14. doi:10.3389/fnagi.2022.791222

37. Fjell AM, Rogeberg O, Sørensen Ø, et al. Reevaluating the role of education on cognitive decline and brain aging in longitudinal cohorts across 33 Western countries. Nat Med. 2025;31(9):2967–2976. doi:10.1038/s41591-025-03828-y

38. Judd N, Kievit R. No effect of additional education on long-term brain structure, a preregistered natural experiment in thousands of individuals. Lerch JP, Behrens TE, eds. eLife. 2025;13:RP101526. doi:10.7554/eLife.101526

39. Lyall DM, Quinn T, Lyall LM, et al. Quantifying bias in psychological and physical health in the UK Biobank imaging sub-sample. Brain Commun. 2022;4(3):fcac119. doi:10.1093/braincomms/fcac119

40. Cox SR, Lyall DM, Ritchie SJ, et al. Associations between vascular risk factors and brain MRI indices in UK Biobank. Eur Heart J. 2019;40(28):2290–2300. doi:10.1093/eurheartj/ehz100

41. Linli Z, Feng J, Zhao W, Guo S. Associations between smoking and accelerated brain ageing. Prog Neuropsychopharmacol Biol Psychiatry. 2022;113:110471. doi:10.1016/j.pnpbp.2021.110471

42. Chen H, Cao Z, Zhang J, Li D, Wang Y, Xu C. Accelerometer-Measured Physical Activity and Neuroimaging-Driven Brain Age. Health Data Sci. 2025;5:0257. doi:10.34133/hds.0257

43. Holtermann A, Krause N, Beek AJ van der, Straker L. The physical activity paradox: six reasons why occupational physical activity (OPA) does not confer the cardiovascular health benefits that leisure time physical activity does. Br J Sports Med. 2018;52(3):149–150. doi:10.1136/bjsports-2017-097965

44. Coenen P, Huysmans MA, Holtermann A, et al. Do highly physically active workers die early? A systematic review with meta-analysis of data from 193 696 participants. Br J Sports Med. 2018;52(20):1320–1326. doi:10.1136/bjsports-2017-098540

45. Elbejjani M, Auer R, Jacobs DR, et al. Cigarette smoking and gray matter brain volumes in middle age adults: the CARDIA Brain MRI sub-study. Transl Psychiatry. 2019;9(1):78. doi:10.1038/s41398-019-0401-1

46. Moore SC, Orpen B, Smith J, et al. Alcohol affordability: implications for alcohol price policies. A cross-sectional analysis in middle and older adults from UK Biobank. J Public Health. 2022;44(2):e192–e202. doi:10.1093/pubmed/fdab095

47. Boyd J, Hayes K, Green D, Angus C, Holmes J. The contribution of health behaviour to socioeconomic inequalities in alcohol harm: Analysis of the UK biobank, a large cohort study with linked health outcomes. SSM - Popul Health. 2023;23:101443. doi:10.1016/j.ssmph.2023.101443

48. Daviet R, Aydogan G, Jagannathan K, et al. Associations between alcohol consumption and gray and white matter volumes in the UK Biobank. Nat Commun. 2022;13(1):1175. doi:10.1038/s41467-022-28735-5

49. Fry A, Littlejohns TJ, Sudlow C, et al. Comparison of Sociodemographic and Health-Related Characteristics of UK Biobank Participants With Those of the General Population. Am J Epidemiol. 2017;186(9):1026–1034. doi:10.1093/aje/kwx246

50. Bontempi D, Benini S, Signoroni A, Svanera M, Muckli L. CEREBRUM: a fast and fully-volumetric Convolutional Encoder-decodeR for weakly-supervised sEgmentation of BRain strUctures from out-of-the-scanner MRI. Med Image Anal. 2020;62:101688. doi:10.1016/j.media.2020.101688

51. Svanera M, Savardi M, Signoroni A, Benini S, Muckli L. Fighting the scanner effect in brain MRI segmentation with a progressive level-of-detail network trained on multi-site data. Med Image Anal. 2024;93:103090. doi:10.1016/j.media.2024.103090

52. Fischl B. FreeSurfer. NeuroImage. 2012;62(2):774–781. doi:10.1016/j.neuroimage.2012.01.021

53. Dalby C, Dibble A, Benini S, et al. NeuroMorph: A Unified Morphological Reference Space for Cross-Disease Brain Profiling. medRxiv. Preprint posted online August 14, 2026:2026.08.13.26359403. doi:10.64898/2026.08.13.26359403

