## Supplement 1 for "Modifiable Contributors to Socioeconomic Inequality in Brain Aging"

### Supplementary

**eMethods 1.** Socioeconomic status coding

**eFigure 1.** Distribution of Household Income and Educational Attainment in the Analytic Sample.

**eMethods 2.** Risk Factor Coding

**eMethods 3**. Brain Age Estimation

**eFigure 2**. Brain Age Prediction Performance

**eMethods 4.** Statistical analyses

**eFigure 3.** Concordance of T1 and T2 BAG Estimates

**eTable1**. Associations of Socioeconomic Indicators With T1-weighted and T2-FLAIR Brain Age Gap

**eFigure 4.** Area deprivation is associated with higher BAG when using IMD as the area-deprivation metric

**eFigure 5**. The divergence in sign for the association between education and T2 BAG is driven by female participants.

**eTable 2.** Interactions Between Educational Attainment and Sex and Age in Associations With Brain Age Gap

**eTable3.** Associations of Socioeconomic Indicators with Individual Risk Factors

**eFigure 6.** Area deprivation is associated with worse CV risk and health behaviors when using IMD as the area-deprivation metric.

**eTable 4.** Associations of Individual Risk Factors with T1-weighted and T2-FLAIR Brain Age Gap

**eFigure 7.** LE8 mediates a greater proportion of the IMD-BAG association than the TDI-BAG association.

**eTable 5.** Mediation by the Life's Essential 8 Score of Associations Between Socioeconomic Indicators and Brain Age Gap

**eTable 6**. Parallel Mediation by Individual Risk Factors of Associations Between Household Income and T1-weighted Brain Age Gap

**eTable 7**. Parallel Mediation by Individual Risk Factors of Associations Between Household Income and T2-FLAIR Brain Age Gap

**eTable 8**. Parallel Mediation by Individual Risk Factors of Associations Between Townsend Deprivation Index and T1-weighted Brain Age Gap

**eTable 9**. Parallel Mediation by Individual Risk Factors of Associations Between Townsend Deprivation Index and T2-FLAIR Brain Age Gap

**eFigure 8**. Smoking carries most of the education→ BAG association.

### eMethods 1. Socioeconomic status coding

*Household income*. Average total pre-tax household income was recorded in five bands, mapped to an ordinal score: 1, less than £18,000; 2, £18,000-30,999; 3, £31,000-51,999; 4, £52,000-100,000; 5, greater than £100,000. "Do not know" and "prefer not to answer" were set to missing. For regression models, the upper two bands were merged, resulting in four categories (< £18,000, 18,000-30,999, £31,000-51,999; 4, ≥ £52,000), which were dummy coded with the highest-income group (≥ £52,000) as the reference category. For mediation models, the full five level ordinal score was entered as a continuous variable.

*Education*. Each participant's educational qualifications were recorded across six fields. The highest qualification across these fields was taken and mapped to a 7-level hierarchy: 1, none of the above; 2, CSEs or equivalent; 3, O levels/GCSEs or equivalent; 4, A levels/AS levels or equivalent; 5, NVG, HND, or HNC or equivalent; 6, other professional qualifications (eg nursing, teaching); and 7, college or university degree. "Prefer not to answer" was set to missing only where no valid qualification was present. For regression models, the 7-level scale was collapsed to four levels: non (1); secondary (2-4); post-secondary (5-6); and degree (7), then dummy coded with degree as the reference category. For mediation models, the 7-level ordinal score was entered as a continuous variable.

*Townsend Deprivation Index.* TDI, an area-based measure of material deprivation, was used as the primary area-level metric. For regression models, TDI was divided into quartiles and dummy coded, with the least deprived quartile (Q1) was the reference category. For mediation models, TDI was z-scored within the analytic sample and entered as a continuous variable.

*Index of Multiple Deprivation.* In sensitivity analyses, the Index of Multiple Deprivation (IMD; England only) replaced TDI as the area level deprivation metric and was handled identically. As IMD is defined differently for each UK constituent country, these analyses were restricted to the England-based subsample.

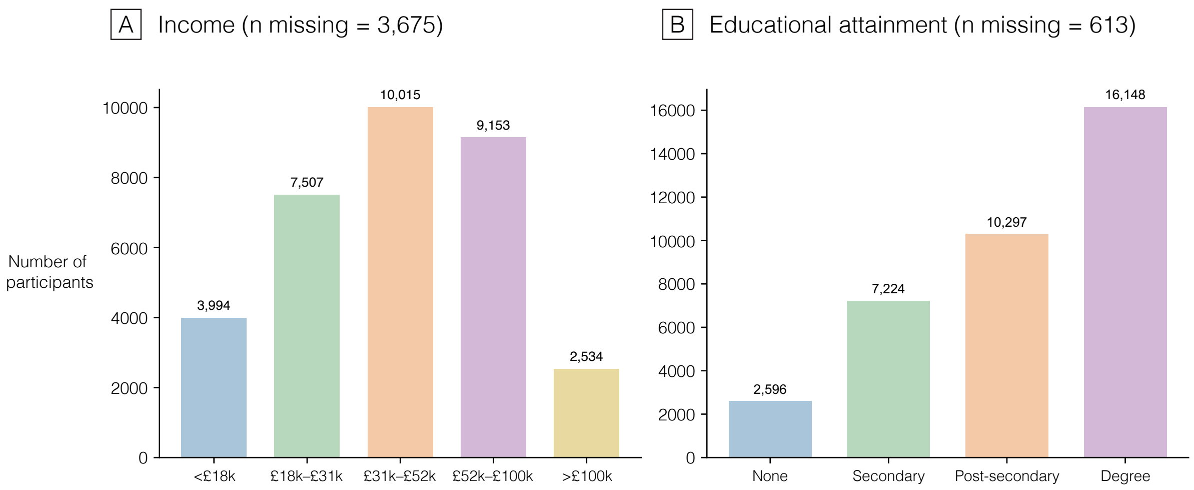


**eFigure 1. Distribution of Household Income and Educational Attainment in the Analytic Sample.** Bars show the number of participants in each category. Household income is annual pre-tax household income; educational attainment is the highest qualification reported. Both distributions are based on the analytic sample of 36 878 participants; counts exclude participants with missing data for the indicator shown, as noted in each panel heading.

### eMethods 2. Risk Factor Coding

*Smoking*. Smoking status was coded as never, previous, or current. Pack years of cigarettes was used as the primary smoking risk factor, with never-smokers set to 0 pack years. Values were Winsorized at the 99$^{th}$ percentile of ever-smokers, then log-transformed and z-scored.

*Alcohol*. Alcohol drinker status was coded as never, previous, or current. Weekly units of alcohol were calculated from reported weekly beverage intake using UK-unit conversions following Topiwala et all[1] and set to 0 for participants reporting 'never' for alcohol frequency. Weekly units were capped at 150. Participants that reported health related reasons for stopping drinking (illness, doctor's advice, health precaution) were set to missing to mitigate sick-quitter bias.

*Body mass index*. BMI was retained as continuous and z-scored. Implausible value (<15 or >60) were set to missing.

*Glycated hemoglobin.* Blood glucose was measured as HbA1c and converted to percentage of glycated hemoglobin, then z-scored.

*Cholesterol.* Non-high-density lipoprotein (HDL) cholesterol was calculated from total cholesterol minus HDL cholesterol, then binarized as high (>= 3.4 mmol/L or on cholesterol lowering medication) or normal (<3.4 mmol/L).

*Blood pressure*. Systolic (SBP) and diastolic (DBP) blood pressure were each computed as a mean of two automated readings, entered as separate variables and z-scored. Implausible values were set to missing.

*Diet*. Diet quality was captured using a 9-component score adapted for UK Biobank [2]. Each component was scored as unhealthy (1) or not (0): red meat (>1×/week), fish (<2×/week), fruit and vegetables (<5 servings/day), processed meat (>1×/week), milk type, spread use, cereal (<5 bowls/week), added salt, and water (<6 glasses/day). Components were summed (0–9) and reverse-coded so that higher values reflect a worse diet, matching the directionality of the other risk factors. The score was entered on its 0–9 scale.

*Physical activity*. Physical activity was calculated as total metabolic-equivalent-of-task (MET) minutes per week, derived from time spent walking and performing moderate and vigorous activity. For analysis, physical inactivity was derived by reverse-coding the activity score, which was then z-scored.

*Sleep*. Sleep duration was self-reported in hours per day and binarized as optimal (7-9 hours) or suboptimal (<7 or >9 hours).

*Social and emotional factors*. Social and emotional risk factors were calculated following Singh et al.,[3]. Stress was converted from frequency feeling tense, fidgety, or restless to a binarized variable representing frequent stress vs no stress. Likewise, infrequent social visits were measured by converting the frequency of family and friend social visits to a binary variable representing frequent (monthly to daily) and infrequent (less than monthly) social visits.

### eMethods 3. Brain Age Estimation

Brain Age was estimated for participants using NeuroFM, a foundation model based on a parameter-efficient 3D convolutional neural network[4]. The model was pretrained by supervised multi-task learning on the LDM100k, a dataset of 100,000 synthetic T1 brain MRI volumes generated by a generative model trained on UK biobank data.

As NeuroFM was pretrained only on T1-weighted data, T2-weighted brain age estimates were obtained by fine-tuning to the T2-weighted contrast. Fine-tuning was performed in previous work[4] using a model development sample of 7,700 UK Biobank participants (15,400 T1 and T2 volumes), selected according to health criteria specified during model development. This sample was defined prior to and independently of the present study, and all 7,700 participants were excluded from all analyses reported here. The remaining 36,878 participants formed the held-out analytic sample. The brain age gap was calculated for each modality as predicted age minus chronological age at imaging (years); where positive values indicate an older-appearing brain. BAG was retained on its raw (years) scale, and all analyses were adjusted for age at imaging.

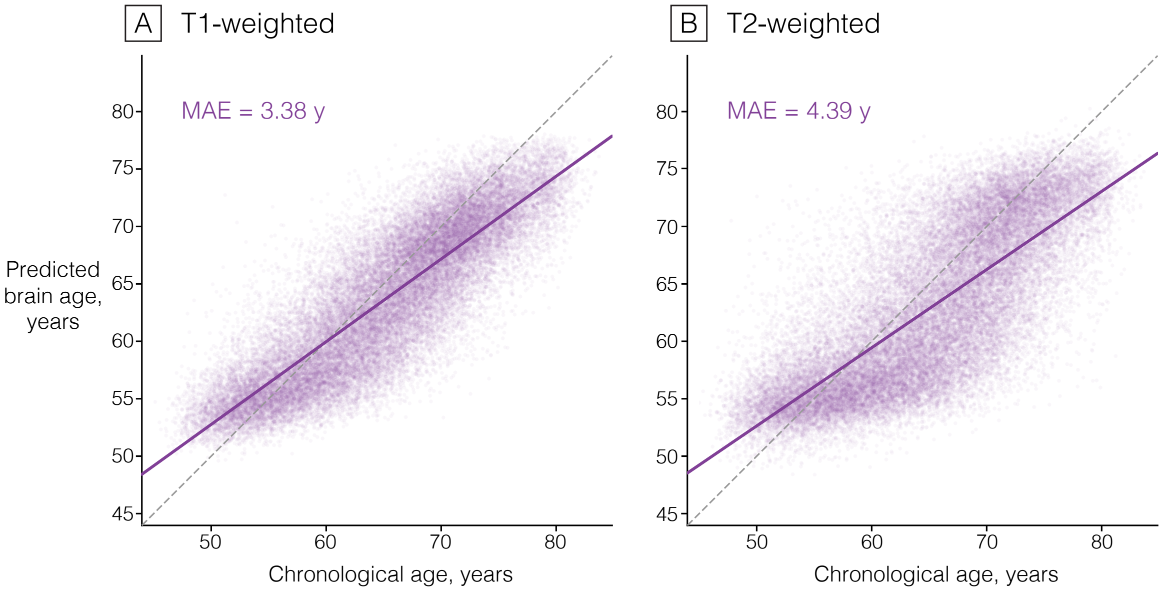


**eFigure 2 | Brain Age Prediction Performance.** Predicted brain age plotted against chronological age for T1-weighted (A) and T2-weighted (B) MRI in the analytic sample (n= 36, 878). Each point represents 1 participant; the solid line is the ordinary least squares fit, and the dashed line is the line of identity, where predicted age = chronological age. Mean absolute error is shown in each panel. For both modalities, the fitted slope was shallower than the line of identity. Brain age predictions were generated with NeuroFM[4].

### eMethods 4. Statistical analyses

Analyses were performed in Python 3.12.3 using statsmodels (0.14.6), pandas (2.3.3), and NumPy (2.4.1).

*Covariates:* For models in which risk factors were the outcome, age at baseline was used as the age covariate. For models in which brain age gap was the outcome, age at imaging was used as the age covariate.

*Regression models.* Each association was estimated with a separate ordinary least squares model fit on complete cases (minimum 1000 per model); 95% CIs were computed as b ± 1.96 * SE. False discovery rate correction (Benjamini-Hochberg) was applied independently within each of three analysis families, polling all exposure-outcome tests within a family but not across families: SES → BAG (2 outcomes, 9 exposure contrasts), risk factor → BAG (2x13), and SES → risk factor (9x13). Income, education, and TDI each contributed three dummy contrasts to the SES families.

*Mediation models.* Mediation followed the three-model structure of Baron and Kenny: model A (Y ~ X + covariates) estimated the total effect c; model B (M ~ X + covariates) estimated path a; and model C (Y ~ X + M + covariates) estimated path b and the direct effect c'. All path models used Heteroskedasticity-consistent (HC3) standard errors and adjusted for age at imaging, sex, and ethnicity. The indirect effect was the product a*b, with 95% Cis from 5000 percentile bootstrap resamples (fixed seed); the proportion mediated was ab/c. Parallel mediation models entered all risk factors simultaneously, yielding mediator-specific indirect effects and a total indirect effect; multicollinearity among mediators was assessed using variance inflation factors on model C. Benjamini-Hochberg FDR correction was applied across the full set of indirect-effect tests.

*Assumptions.* Interpretation of indirect effects as mediation assumes no unmeasured exposure-outcome, mediator-outcome, or exposure-outcome confounding, and correct directional ordering of exposure, mediator, and outcome. Because SES and risk factors were assessed at baseline visit, the exposure-mediator ordering is not established by design. Estimates are therefore interpreted as statistical associations.

*Sensitivity analyses*

*Education-BAG interaction.* Because education was represented by three dummy contrasts, effect modification was assessed as a joint test of all three education moderator interaction terms (3 df Wald test) rather than as contrast-wise tests. Models regressed BAG on education, the moderator, and their interaction, and covariates (age at imaging, sex, ethnicity; sex omitted where it was the moderator). Moderators were sex and age band (≥66 vs <66 years at imaging). These analyses were post hoc and were not included in the FDR families.

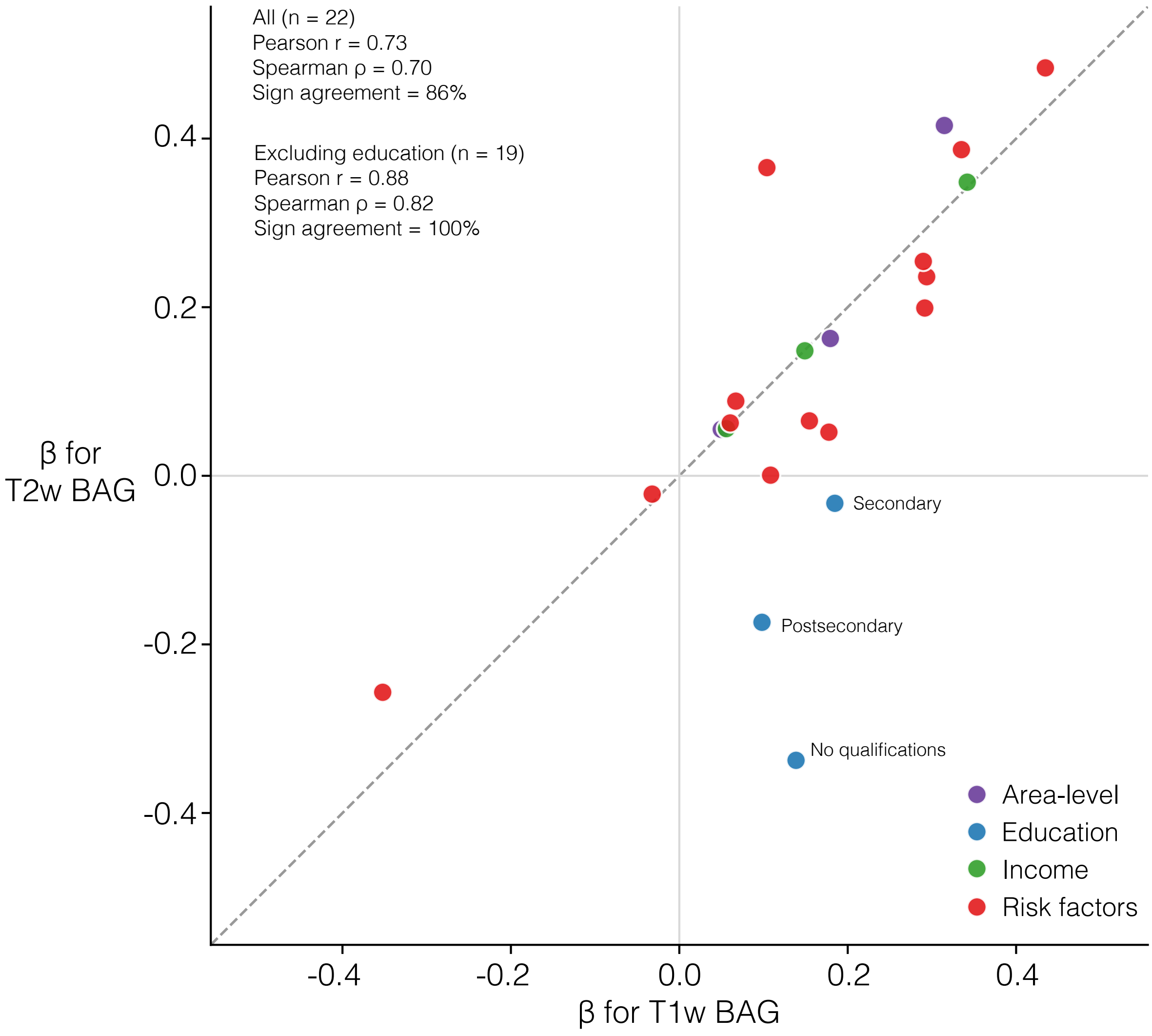


**eFigure 3 | Concordance of T1 and T2 BAG Estimates.** Standardized regression coefficients (β, years of brain age gap per 1-SD higher exposure or per category contrast) for T2-derived brain age gap plotted against the corresponding coefficients for T1-derived brain age gap. Each point represents 1 exposure. Estimates are from separate linear regression models, each fitted with a single exposure and adjusted for age, sex, and ethnicity (22 models per modality: 3 area-level deprivation, 3 education, 3 income, and 13 cardiovascular and lifestyle risk factors). The dashed line is the line of identity. Across all 22 associations, agreement between modalities wash high (Perason r - .73; Spearman ρ = .70; 19 of 22[86%] concordant in sign). The 3 education contrasts were the only associations to differ in sign between modalities, being positive for T1 brain age gap and null to negative for T2 brain age gap. Excluding these contrasts, agreement was stronger (n = 19; r = .88; ρ = .82; 19 of 19[100%] concordant in sign). BAG, brain age gap; T1, T1-weighted; T2, T2-weighted.

### eTable1. Associations of Socioeconomic Indicators With T1-weighted and T2-FLAIR Brain Age Gap [a]

| Exposure | Outcome | No. | β (95% CI) [b] | P | P (FDR) [c] |
|---|---|---|---|---|---|
| **Income <18k** | Brain age gap (T1) | 15,647 | 0.348 (0.223 to 0.472) | <.001 | <.001 |
| **Income 18-31k** | Brain age gap (T1) | 19,152 | 0.156 (0.055 to 0.256) | .002 | .005 |
| **Income 31-52k** | Brain age gap (T1) | 21,662 | 0.059 (-0.029 to 0.147) | .19 | .25 |
| **No Educational Qualifications** | Brain age gap (T1) | 18,686 | 0.139 (-0.000 to 0.278) | .05 | .07 |
| **Secondary Education** | Brain age gap (T1) | 23,303 | 0.184 (0.092 to 0.276) | <.001 | <.001 |
| **Postsecondary Education** | Brain age gap (T1) | 26,372 | 0.094 (0.013 to 0.175) | .02 | .04 |
| **TDI Q4 (most deprived)** | Brain age gap (T1) | 18,180 | 0.329 (0.232 to 0.426) | <.001 | <.001 |
| **TDI Q3** | Brain age gap (T1) | 18,287 | 0.182 (0.086 to 0.278) | <.001 | <.001 |
| **TDI Q2** | Brain age gap (T1) | 18,306 | 0.051 (-0.044 to 0.146) | .29 | .35 |
| **Income <18k** | Brain age gap (T2) | 15,647 | 0.351 (0.187 to 0.515) | <.001 | <.001 |
| **Income 18-31k** | Brain age gap (T2) | 19,152 | 0.155 (0.022 to 0.287) | .02 | .04 |
| **Income 31-52k** | Brain age gap (T2) | 21,662 | 0.056 (-0.060 to 0.173) | .34 | .38 |
| **No Educational Qualifications** | Brain age gap (T2) | 18,686 | -0.333 (-0.519 to -0.148) | <.001 | .001 |
| **Secondary Education** | Brain age gap (T2) | 23,303 | -0.027 (-0.148 to 0.094) | .66 | .66 |
| **Postsecondary Education** | Brain age gap (T2) | 26,372 | -0.173 (-0.281 to -0.066) | .002 | .004 |
| **TDI Q4 (most deprived)** | Brain age gap (T2) | 18,180 | 0.427 (0.300 to 0.554) | <.001 | <.001 |
| **TDI Q3** | Brain age gap (T2) | 18,287 | 0.165 (0.038 to 0.291) | .01 | .02 |
| **TDI Q2** | Brain age gap (T2) | 18,306 | 0.060 (-0.067 to 0.186) | .36 | .38 |

Abbreviations. BAG, brain age gap; CI, confidence interval; FDR, false discovery rate; FLAIR, fluid-attenuated recovery; TDI, Townsend Deprivation Index.

[a] Each row is a separate linear regression model comparing the listed category with its reference category; No. is the combined number of participants in the two categories. Reference categories were household income ≥ £52 000/y, university degree, and TDI quartile 1 (least deprived).

[b] β represents the difference in brain age gap, in years, relative to the reference category. Models were adjusted for age, sex, and ethnicity.

[c] P values were adjusted for multiple comparisons using the Benjamini-Hochberg false discovery rate across the 18 tests reported in this table.

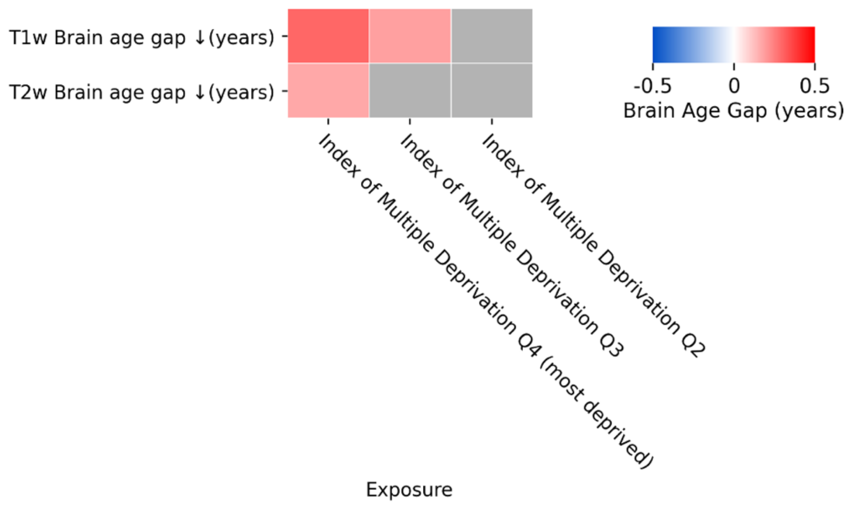


**eFigure 4 | Area deprivation is associated with higher BAG when using IMD as the area-deprivation metric.** Heatmap of regression coefficients from independent OLS models of BAG on IMD, adjusted for age and sex. The results for IMD are on the same scale as for TDI. Rows are outcomes (T1 and T2 BAG). Columns are IMD quartiles, which are dummy coded contrasts against the high-SES reference category (IMD Q1). Cell colors indicate the b (BAG, years) for each model; red indicates a higher BAG (older-appearing brain) and blue indicates lower BAG (younger-appearing brain) relative to the reference category. Each cell is one independent regression. Grey cells are non-significant after Benjamini-Hochberg FDR correction applied across all tests in the figure. BAG, brain age gap; FDR; false discovery rate; IMD, index of multiple deprivation; OLS, ordinary least squares; SES, socioeconomic status; TDI, Townsend Deprivation Index.

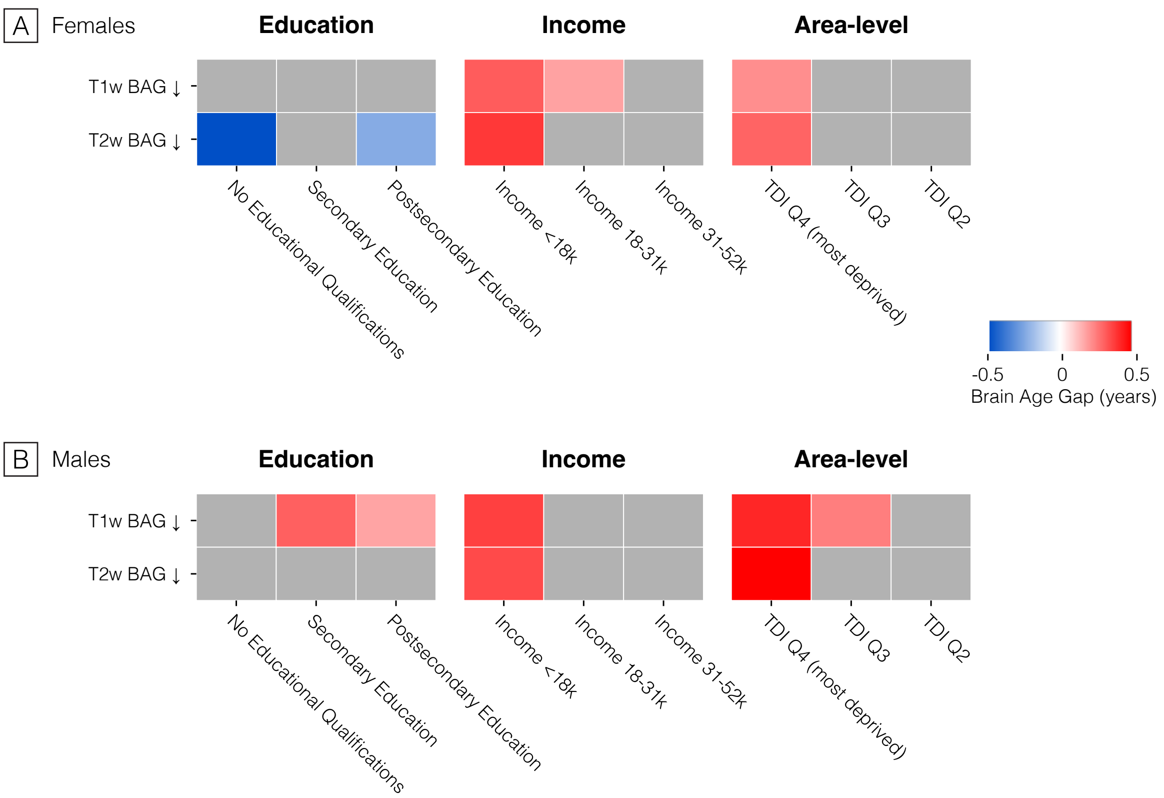


**eFigure 5 | The divergence in sign for the association between education and T2 BAG is driven by female participants.** Heatmaps of regression coefficients from independent OLS models of BAG on each SES indicator, for (**A**) females, and (**B**) males, adjusted for age. Rows are outcomes (T1 and T2 BAG). Columns are SES exposures, where categorical SES indicators (education levels, income brackets, TDI quartiles) are dummy coded contrasts against the high-SES reference category (university degree; income > £52,000; TDI Q1). Cell colors indicate the b (BAG, years) for each model; red indicates a higher BAG (older-appearing brain) and blue indicates lower BAG (younger-appearing brain) relative to the reference category. Each cell is one independent regression. Grey cells are non-significant after Benjamini-Hochberg FDR correction applied across all tests in the figure. BAG, brain age gap; FDR; false discovery rate; OLS, ordinary least squares; SES, socioeconomic status; TDI, Townsend Deprivation Index

### eTable 2. Interactions Between Educational Attainment and Sex and Age in Associations With Brain Age Gap [a]

| Modality | Moderator | Education level [b] | β (95% CI) [c] | P value |
|---|---|---|---|---|
| T1 | Female sex | Joint interaction test | $F_{3,36163}$ = 2.05 | .105 |
| | | No qualifications | -0.040 [-0.319 to 0.239] | .780 |
| | | Secondary | -0.213 [-0.402 to -0.023] | .028 |
| | | Post-secondary | -0.145 [-0.308 to 0.017] | .080 |
| T1 | Age ≥66 y | Joint interaction test | $F_{3,36162}$ = 1.26 | .287 |
| | | No qualifications | -0.324 [-0.661 to 0.014] | .060 |
| | | Secondary | 0.007 [-0.179 to 0.194] | .937 |
| | | Post-secondary | -0.042 [-0.204 to 0.120] | .611 |
| T1 | Female sex × age ≥66 y | Joint interaction test | $F_{3,36161}$ = 2.21 | .085 |
| | | No qualifications | -0.338 [-0.634 to -0.041] | .026 |
| | | Secondary | -0.179 [-0.398 to 0.039] | .107 |
| | | Post-secondary | -0.157 [-0.357 to 0.044] | .126 |
| T2 | Female sex | Joint interaction test | $F_{3,36163}$ = 4.45 | .004 |
| | | No qualifications | -0.582 [-0.949 to -0.214] | .002 |
| | | Secondary | -0.320 [-0.571 to -0.070] | .012 |
| | | Post-secondary | -0.158 [-0.374 to 0.058] | .151 |
| T2 | Age ≥66 y | Joint interaction test | $F_{3,36162}$ = 0.98 | .399 |
| | | No qualifications | -0.208 [-0.624 to 0.209] | .328 |
| | | Secondary | -0.103 [-0.347 to 0.140] | .407 |
| | | Post-secondary | -0.172 [-0.388 to 0.044] | .118 |
| T2 | Female sex × age ≥66 y | Joint interaction test | $F_{3,36161}$ = 5.15 | .001 |
| | | No qualifications | -0.658 [-1.055 to -0.262] | .001 |
| | | Secondary | -0.409 [-0.698 to -0.120] | .006 |
| | | Post-secondary | -0.327 [-0.601 to -0.052] | .020 |

Abbreviations: Bag, brain age gap; CI, confidence interval; FLAIR, fluid-attenuated inversion recovery.

[a] Analyses included 36 173 participants with complete data on education, sex, age, and ethnicity. Models were adjusted for age, sex, and ethnicity

[b] The joint interaction test is an F test of the 3 education x moderator product terms considered simultaneously. Rows below it give the individual product-term coefficients for each education level relative to university education.

[c] β represents the difference in the education-BAG association, in years, associated with the moderator.

### eTable3 | Associations of Socioeconomic Indicators with Individual Risk Factors [a]

| Exposure | Outcome | No. | β (95% CI) [b] | P | P (FDR) [c] |
|---|---|---|---|---|---|
| **Income <18k** | Life's Essential 8 score | 11,841 | -0.264 (-0.305 to -0.223) | <.001 | <.001 |
| **Income 18-31k** | Life's Essential 8 score | 14,489 | -0.143 (-0.175 to -0.110) | <.001 | <.001 |
| **Income 31-52k** | Life's Essential 8 score | 16,362 | -0.124 (-0.153 to -0.095) | <.001 | <.001 |
| **No Educational Qualifications** | Life's Essential 8 score | 13,958 | -0.437 (-0.483 to -0.391) | <.001 | <.001 |
| **Secondary Education** | Life's Essential 8 score | 17,523 | -0.274 (-0.303 to -0.244) | <.001 | <.001 |
| **Postsecondary Education** | Life's Essential 8 score | 19,900 | -0.219 (-0.246 to -0.193) | <.001 | <.001 |
| **TDI Q4 (most deprived)** | Life's Essential 8 score | 13,417 | -0.190 (-0.222 to -0.159) | <.001 | <.001 |
| **TDI Q3** | Life's Essential 8 score | 13,607 | -0.096 (-0.127 to -0.065) | <.001 | <.001 |
| **TDI Q2** | Life's Essential 8 score | 13,533 | -0.038 (-0.069 to -0.007) | .02 | .02 |
| **Income <18k** | BMI (z) | 15,624 | 0.163 (0.126 to 0.201) | <.001 | <.001 |
| **Income 18-31k** | BMI (z) | 19,123 | 0.128 (0.098 to 0.158) | <.001 | <.001 |
| **Income 31-52k** | BMI (z) | 21,637 | 0.092 (0.066 to 0.119) | <.001 | <.001 |
| **No Educational Qualifications** | BMI (z) | 18,659 | 0.299 (0.258 to 0.341) | <.001 | <.001 |
| **Secondary Education** | BMI (z) | 23,269 | 0.166 (0.139 to 0.193) | <.001 | <.001 |
| **Postsecondary Education** | BMI (z) | 26,335 | 0.198 (0.173 to 0.222) | <.001 | <.001 |
| **TDI Q4 (most deprived)** | BMI (z) | 18,152 | 0.115 (0.086 to 0.145) | <.001 | <.001 |
| **TDI Q3** | BMI (z) | 18,263 | 0.088 (0.060 to 0.116) | <.001 | <.001 |
| **TDI Q2** | BMI (z) | 18,278 | 0.040 (0.013 to 0.068) | .004 | .006 |
| **Income <18k** | Diet risk score ↓(z) | 15,647 | 0.175 (0.121 to 0.229) | <.001 | <.001 |
| **Income 18-31k** | Diet risk score ↓(z) | 19,152 | 0.122 (0.079 to 0.165) | <.001 | <.001 |
| **Income 31-52k** | Diet risk score ↓(z) | 21,662 | 0.123 (0.085 to 0.161) | <.001 | <.001 |
| **No Educational Qualifications** | Diet risk score ↓(z) | 18,686 | 0.265 (0.205 to 0.325) | <.001 | <.001 |
| **Secondary Education** | Diet risk score ↓(z) | 23,303 | 0.202 (0.163 to 0.242) | <.001 | <.001 |
| **Postsecondary Education** | Diet risk score ↓(z) | 26,372 | 0.143 (0.109 to 0.178) | <.001 | <.001 |
| **TDI Q4 (most deprived)** | Diet risk score ↓(z) | 18,180 | 0.158 (0.116 to 0.200) | <.001 | <.001 |
| **TDI Q3** | Diet risk score ↓(z) | 18,287 | 0.097 (0.056 to 0.137) | <.001 | <.001 |
| **TDI Q2** | Diet risk score ↓(z) | 18,306 | 0.030 (-0.011 to 0.070) | .15 | .18 |
| **Income <18k** | HbA1c (%) (z) | 14,717 | 0.142 (0.103 to 0.182) | <.001 | <.001 |
| **Income 18-31k** | HbA1c (%) (z) | 18,026 | 0.093 (0.061 to 0.124) | <.001 | <.001 |
| **Income 31-52k** | HbA1c (%) (z) | 20,327 | 0.055 (0.028 to 0.083) | <.001 | <.001 |
| **No Educational Qualifications** | HbA1c (%) (z) | 17,504 | 0.103 (0.061 to 0.146) | <.001 | <.001 |
| **Secondary Education** | HbA1c (%) (z) | 21,847 | 0.070 (0.042 to 0.098) | <.001 | <.001 |
| **Postsecondary Education** | HbA1c (%) (z) | 24,780 | 0.074 (0.048 to 0.099) | <.001 | <.001 |
| **TDI Q4 (most deprived)** | HbA1c (%) (z) | 16,797 | 0.083 (0.053 to 0.114) | <.001 | <.001 |
| **TDI Q3** | HbA1c (%) (z) | 16,949 | 0.068 (0.039 to 0.098) | <.001 | <.001 |
| **TDI Q2** | HbA1c (%) (z) | 16,981 | 0.025 (-0.003 to 0.054) | .08 | .10 |
| **Income <18k** | Pack years smoking (z) | 13,382 | 0.294 (0.252 to 0.336) | <.001 | <.001 |
| **Income 18-31k** | Pack years smoking (z) | 16,380 | 0.193 (0.159 to 0.227) | <.001 | <.001 |
| **Income 31-52k** | Pack years smoking (z) | 18,538 | 0.110 (0.080 to 0.139) | <.001 | <.001 |
| **No Educational Qualifications** | Pack years smoking (z) | 15,892 | 0.460 (0.415 to 0.506) | <.001 | <.001 |
| **Secondary Education** | Pack years smoking (z) | 19,931 | 0.235 (0.205 to 0.265) | <.001 | <.001 |
| **Postsecondary Education** | Pack years smoking (z) | 22,526 | 0.207 (0.180 to 0.234) | <.001 | <.001 |
| **TDI Q4 (most deprived)** | Pack years smoking (z) | 15,481 | 0.341 (0.308 to 0.375) | <.001 | <.001 |
| **TDI Q3** | Pack years smoking (z) | 15,640 | 0.143 (0.111 to 0.174) | <.001 | <.001 |
| **TDI Q2** | Pack years smoking (z) | 15,651 | 0.049 (0.018 to 0.080) | .002 | .003 |
| **Income <18k** | High non-HDL cholesterol | 15,647 | 0.027 (0.012 to 0.043) | <.001 | <.001 |
| **Income 18-31k** | High non-HDL cholesterol | 19,152 | 0.020 (0.007 to 0.032) | .002 | .003 |
| **Income 31-52k** | High non-HDL cholesterol | 21,662 | 0.009 (-0.002 to 0.020) | .12 | .15 |
| **No Educational Qualifications** | High non-HDL cholesterol | 18,686 | 0.035 (0.017 to 0.052) | <.001 | <.001 |
| **Secondary Education** | High non-HDL cholesterol | 23,303 | 0.026 (0.015 to 0.038) | <.001 | <.001 |

### eTable3 | continued

| Exposure | Outcome | No. | β (95% CI) [b] | P | P (FDR) [c] |
|---|---|---|---|---|---|
| **Postsecondary Education** | High non-HDL cholesterol | 26,372 | 0.027 (0.017 to 0.037) | <.001 | <.001 |
| **TDI Q4 (most deprived)** | High non-HDL cholesterol | 18,180 | 0.006 (-0.006 to 0.018) | .35 | .41 |
| **TDI Q3** | High non-HDL cholesterol | 18,287 | -0.001 (-0.013 to 0.010) | .82 | .84 |
| **TDI Q2** | High non-HDL cholesterol | 18,306 | -0.010 (-0.021 to 0.002) | .11 | .14 |
| **Income <18k** | Diastolic blood pressure ↓(z) | 14,732 | -0.007 (-0.045 to 0.031) | .71 | .75 |
| **Income 18-31k** | Diastolic blood pressure ↓(z) | 18,036 | -0.008 (-0.039 to 0.022) | .59 | .65 |
| **Income 31-52k** | Diastolic blood pressure ↓(z) | 20,398 | 0.036 (0.009 to 0.063) | .009 | .01 |
| **No Educational Qualifications** | Diastolic blood pressure ↓(z) | 17,536 | 0.078 (0.036 to 0.121) | <.001 | <.001 |
| **Secondary Education** | Diastolic blood pressure ↓(z) | 21,960 | 0.083 (0.055 to 0.111) | <.001 | <.001 |
| **Postsecondary Education** | Diastolic blood pressure ↓(z) | 24,859 | 0.067 (0.042 to 0.091) | <.001 | <.001 |
| **TDI Q4 (most deprived)** | Diastolic blood pressure ↓(z) | 17,026 | -0.035 (-0.064 to -0.005) | .02 | .03 |
| **TDI Q3** | Diastolic blood pressure ↓(z) | 17,152 | -0.006 (-0.035 to 0.022) | .66 | .70 |
| **TDI Q2** | Diastolic blood pressure ↓(z) | 17,143 | -0.007 (-0.036 to 0.022) | .63 | .69 |
| **Income <18k** | Systolic blood pressure (z) | 14,732 | 0.066 (0.030 to 0.102) | <.001 | <.001 |
| **Income 18-31k** | Systolic blood pressure (z) | 18,036 | 0.053 (0.024 to 0.082) | <.001 | <.001 |
| **Income 31-52k** | Systolic blood pressure (z) | 20,398 | 0.079 (0.053 to 0.104) | <.001 | <.001 |
| **No Educational Qualifications** | Systolic blood pressure (z) | 17,536 | 0.176 (0.135 to 0.217) | <.001 | <.001 |
| **Secondary Education** | Systolic blood pressure (z) | 21,960 | 0.109 (0.082 to 0.136) | <.001 | <.001 |
| **Postsecondary Education** | Systolic blood pressure (z) | 24,859 | 0.086 (0.063 to 0.110) | <.001 | <.001 |
| **TDI Q4 (most deprived)** | Systolic blood pressure (z) | 17,026 | -0.032 (-0.060 to -0.004) | .03 | .04 |
| **TDI Q3** | Systolic blood pressure (z) | 17,152 | -0.008 (-0.036 to 0.020) | .57 | .64 |
| **TDI Q2** | Systolic blood pressure (z) | 17,143 | -0.002 (-0.030 to 0.026) | .89 | .90 |
| **Income <18k** | Suboptimal sleep | 15,623 | 0.030 (0.014 to 0.046) | <.001 | <.001 |
| **Income 18-31k** | Suboptimal sleep | 19,125 | 0.005 (-0.008 to 0.018) | .48 | .54 |
| **Income 31-52k** | Suboptimal sleep | 21,644 | -0.001 (-0.012 to 0.011) | .93 | .93 |
| **No Educational Qualifications** | Suboptimal sleep | 18,651 | 0.075 (0.057 to 0.092) | <.001 | <.001 |
| **Secondary Education** | Suboptimal sleep | 23,262 | 0.032 (0.020 to 0.043) | <.001 | <.001 |
| **Postsecondary Education** | Suboptimal sleep | 26,325 | 0.029 (0.019 to 0.039) | <.001 | <.001 |
| **TDI Q4 (most deprived)** | Suboptimal sleep | 18,128 | 0.049 (0.037 to 0.062) | <.001 | <.001 |
| **TDI Q3** | Suboptimal sleep | 18,245 | 0.021 (0.009 to 0.033) | <.001 | <.001 |
| **TDI Q2** | Suboptimal sleep | 18,274 | 0.011 (-0.001 to 0.023) | .07 | .09 |
| **Income <18k** | Physical inactivity (z) | 15,426 | -0.244 (-0.280 to -0.209) | <.001 | <.001 |
| **Income 18-31k** | Physical inactivity (z) | 18,875 | -0.222 (-0.252 to -0.193) | <.001 | <.001 |
| **Income 31-52k** | Physical inactivity (z) | 21,394 | -0.107 (-0.132 to -0.083) | <.001 | <.001 |
| **No Educational Qualifications** | Physical inactivity (z) | 18,360 | -0.315 (-0.354 to -0.277) | <.001 | <.001 |
| **Secondary Education** | Physical inactivity (z) | 22,967 | -0.105 (-0.130 to -0.080) | <.001 | <.001 |
| **Postsecondary Education** | Physical inactivity (z) | 25,942 | -0.171 (-0.194 to -0.148) | <.001 | <.001 |
| **TDI Q4 (most deprived)** | Physical inactivity (z) | 17,658 | -0.068 (-0.097 to -0.039) | <.001 | <.001 |
| **TDI Q3** | Physical inactivity (z) | 17,785 | -0.054 (-0.082 to -0.026) | <.001 | <.001 |
| **TDI Q2** | Physical inactivity (z) | 17,802 | -0.020 (-0.048 to 0.009) | .17 | .21 |
| **Income <18k** | Alcohol units weekly (z) | 12,934 | -0.098 (-0.143 to -0.054) | <.001 | <.001 |
| **Income 18-31k** | Alcohol units weekly (z) | 15,906 | -0.107 (-0.140 to -0.073) | <.001 | <.001 |
| **Income 31-52k** | Alcohol units weekly (z) | 18,262 | -0.054 (-0.083 to -0.025) | <.001 | <.001 |
| **No Educational Qualifications** | Alcohol units weekly (z) | 15,369 | 0.013 (-0.033 to 0.060) | .57 | .64 |
| **Secondary Education** | Alcohol units weekly (z) | 19,065 | 0.057 (0.027 to 0.087) | <.001 | <.001 |

### eTable3 | continued

| Exposure | Outcome | No. | β (95% CI) [b] | P | P (FDR) [c] |
|---|---|---|---|---|---|
| **Postsecondary Education** | Alcohol units weekly (z) | 21,531 | 0.007 (-0.019 to 0.034) | .58 | .64 |
| **TDI Q4 (most deprived)** | Alcohol units weekly (z) | 14,565 | 0.119 (0.086 to 0.152) | <.001 | <.001 |
| **TDI Q3** | Alcohol units weekly (z) | 14,967 | 0.046 (0.015 to 0.077) | .003 | .005 |
| **TDI Q2** | Alcohol units weekly (z) | 15,138 | -0.007 (-0.036 to 0.023) | .66 | .70 |
| **Income <18k** | Infrequent social visits | 15,622 | 0.001 (-0.009 to 0.012) | .84 | .86 |
| **Income 18-31k** | Infrequent social visits | 19,132 | -0.012 (-0.020 to -0.004) | .005 | .007 |
| **Income 31-52k** | Infrequent social visits | 21,642 | -0.011 (-0.018 to -0.004) | .003 | .004 |
| **No Educational Qualifications** | Infrequent social visits | 18,640 | -0.008 (-0.020 to 0.003) | .15 | .19 |
| **Secondary Education** | Infrequent social visits | 23,261 | -0.015 (-0.023 to -0.008) | <.001 | <.001 |
| **Postsecondary Education** | Infrequent social visits | 26,322 | -0.014 (-0.021 to -0.008) | <.001 | <.001 |
| **TDI Q4 (most deprived)** | Infrequent social visits | 17,899 | 0.018 (0.010 to 0.026) | <.001 | <.001 |
| **TDI Q3** | Infrequent social visits | 17,996 | -0.001 (-0.008 to 0.006) | .77 | .80 |
| **TDI Q2** | Infrequent social visits | 18,030 | -0.001 (-0.009 to 0.006) | .73 | .76 |
| **Income <18k** | Stress (tense/restless) | 15,346 | 0.061 (0.044 to 0.077) | <.001 | <.001 |
| **Income 18-31k** | Stress (tense/restless) | 18,803 | 0.042 (0.029 to 0.056) | <.001 | <.001 |
| **Income 31-52k** | Stress (tense/restless) | 21,292 | 0.019 (0.007 to 0.031) | .001 | .002 |
| **No Educational Qualifications** | Stress (tense/restless) | 18,309 | 0.007 (-0.011 to 0.026) | .44 | .51 |
| **Secondary Education** | Stress (tense/restless) | 22,831 | 0.005 (-0.008 to 0.017) | .46 | .53 |
| **Postsecondary Education** | Stress (tense/restless) | 25,815 | -0.004 (-0.015 to 0.006) | .43 | .51 |
| **TDI Q4 (most deprived)** | Stress (tense/restless) | 17,717 | 0.034 (0.021 to 0.047) | <.001 | <.001 |
| **TDI Q3** | Stress (tense/restless) | 17,881 | 0.010 (-0.003 to 0.022) | .12 | .16 |
| **TDI Q2** | Stress (tense/restless) | 17,916 | 0.007 (-0.006 to 0.019) | .29 | .35 |

Abbreviations. CI, confidence interval; FDR, false discovery rate; TDI, Townsend Deprivation Index.

[a] Each row is a separate linear regression model comparing the listed category with its reference category; No. is the combined number of participants in the two categories. Reference categories were household income ≥ £52 000/y, university degree, and TDI quartile 1 (least deprived).

[b] β represents the standardized difference in risk factor relative to the reference category. Models were adjusted for age, sex, and ethnicity.

[c] P values were adjusted for multiple comparisons using the Benjamini-Hochberg false discovery rate across the 66 tests reported in this table.

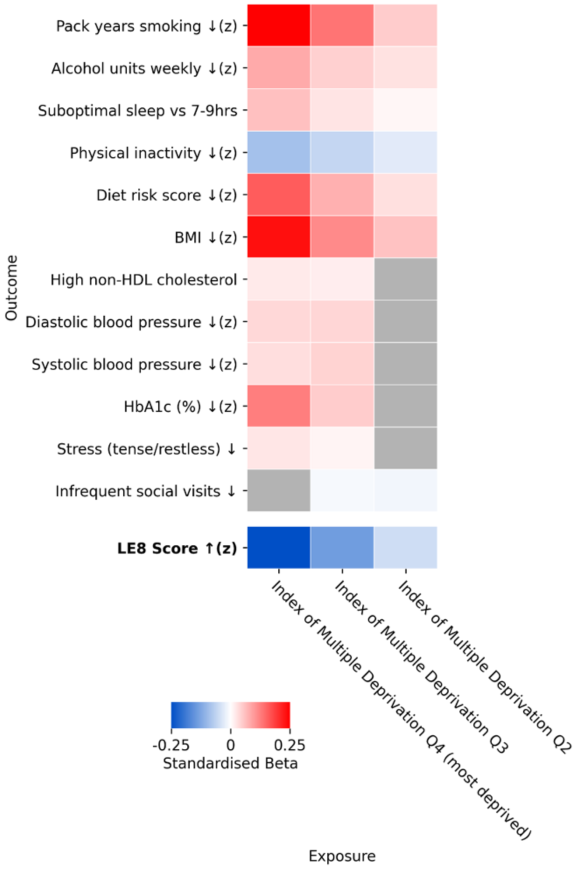


**eFigure 6 | Area deprivation is associated with worse CV risk and health behaviors when using IMD as the area-deprivation metric.** Heatmap of regression coefficients from independent OLS models of risk factors on IMD, adjusted for age and sex. The results for IMD are on the same scale as for TDI. Notably, the associations between high area deprivation and high cholesterol and blood pressure are significant when employing IMD as the area-deprivation metric. Rows are outcomes (T1 and T2 BAG). Columns are IMD quartiles, which are dummy coded contrasts against the high-SES reference category (IMD Q1). Cell colors indicate the b (BAG, years) for each model; red indicates a higher BAG (older-appearing brain) and blue indicates lower BAG (younger-appearing brain) relative to the reference category. Each cell is one independent regression. Grey cells are non-significant after Benjamini-Hochberg FDR correction applied across all tests in the figure. BAG, brain age gap; FDR; false discovery rate; IMD, index of multiple deprivation; OLS, ordinary least squares; SES, socioeconomic status; TDI, Townsend Deprivation Index

### eTable 4 | Associations of Individual Risk Factors with T1-weighted and T2-FLAIR Brain Age Gap [a]

| Exposure | Outcome | No. | β (95% CI) [b] | P | P (FDR) [c] |
|---|---|---|---|---|---|
| **Life's Essential 8 score** | Brain age gap (T1) | 27,259 | -0.354 (-0.395 to -0.312) | <.001 | <.001 |
| **BMI (z)** | Brain age gap (T1) | 36,718 | 0.154 (0.120 to 0.188) | <.001 | <.001 |
| **Diet risk score ↓(z)** | Brain age gap (T1) | 36,772 | 0.058 (0.034 to 0.082) | <.001 | <.001 |
| **HbA1c (%) (z)** | Brain age gap (T1) | 34,067 | 0.295 (0.260 to 0.330) | <.001 | <.001 |
| **Pack years smoking (z)** | Brain age gap (T1) | 31,371 | 0.335 (0.300 to 0.370) | <.001 | <.001 |
| **High non-HDL cholesterol** | Brain age gap (T1) | 36,772 | 0.109 (0.026 to 0.192) | .01 | .01 |
| **Diastolic blood pressure ↓(z)** | Brain age gap (T1) | 34,514 | 0.293 (0.258 to 0.329) | <.001 | <.001 |
| **Systolic blood pressure (z)** | Brain age gap (T1) | 34,514 | 0.291 (0.254 to 0.328) | <.001 | <.001 |
| **Suboptimal sleep** | Brain age gap (T1) | 36,686 | 0.186 (0.105 to 0.266) | <.001 | <.001 |
| **Physical inactivity (z)** | Brain age gap (T1) | 35,738 | -0.031 (-0.066 to 0.004) | .08 | .10 |
| **Alcohol units weekly (z)** | Brain age gap (T1) | 29,627 | 0.434 (0.396 to 0.472) | <.001 | <.001 |
| **Infrequent social visits** | Brain age gap (T1) | 36,186 | 0.111 (-0.019 to 0.241) | .09 | .12 |
| **Stress (tense/restless)** | Brain age gap (T1) | 35,893 | 0.066 (-0.014 to 0.146) | .11 | .13 |
| **Life's Essential 8 score** | Brain age gap (T2) | 27,259 | -0.258 (-0.313 to -0.203) | <.001 | <.001 |
| **BMI (z)** | Brain age gap (T2) | 36,718 | 0.064 (0.020 to 0.109) | .005 | .007 |
| **Diet risk score ↓(z)** | Brain age gap (T2) | 36,772 | 0.061 (0.029 to 0.092) | <.001 | <.001 |
| **HbA1c (%) (z)** | Brain age gap (T2) | 34,067 | 0.259 (0.213 to 0.305) | <.001 | <.001 |
| **Pack years smoking (z)** | Brain age gap (T2) | 31,371 | 0.388 (0.342 to 0.433) | <.001 | <.001 |
| **High non-HDL cholesterol** | Brain age gap (T2) | 36,772 | 0.002 (-0.107 to 0.112) | .97 | .97 |
| **Diastolic blood pressure ↓(z)** | Brain age gap (T2) | 34,514 | 0.236 (0.188 to 0.283) | <.001 | <.001 |
| **Systolic blood pressure (z)** | Brain age gap (T2) | 34,514 | 0.198 (0.150 to 0.247) | <.001 | <.001 |
| **Suboptimal sleep** | Brain age gap (T2) | 36,686 | 0.058 (-0.048 to 0.165) | .28 | .30 |
| **Physical inactivity (z)** | Brain age gap (T2) | 35,738 | -0.022 (-0.068 to 0.024) | .35 | .36 |
| **Alcohol units weekly (z)** | Brain age gap (T2) | 29,627 | 0.485 (0.434 to 0.535) | <.001 | <.001 |
| **Infrequent social visits** | Brain age gap (T2) | 36,186 | 0.376 (0.204 to 0.547) | <.001 | <.001 |
| **Stress (tense/restless)** | Brain age gap (T2) | 35,893 | 0.085 (-0.020 to 0.190) | .11 | .13 |

Abbreviations. BAG, brain age gap; CI, confidence interval; FDR, false discovery rate; FLAIR, fluid-attenuated recovery; TDI, Townsend Deprivation Index.

[a] Each row is a separate linear regression model comparing the listed category with its reference category; No. is the combined number of participants in the two categories. Reference categories were household income ≥ £52 000/y, university degree, and TDI quartile 1 (least deprived).

[b] β represents the difference in brain age gap, in years, per 1 SD difference in risk factor. Models were adjusted for age, sex, and ethnicity.

[c] P values were adjusted for multiple comparisons using the Benjamini-Hochberg false discovery rate across the 26 tests reported in this table

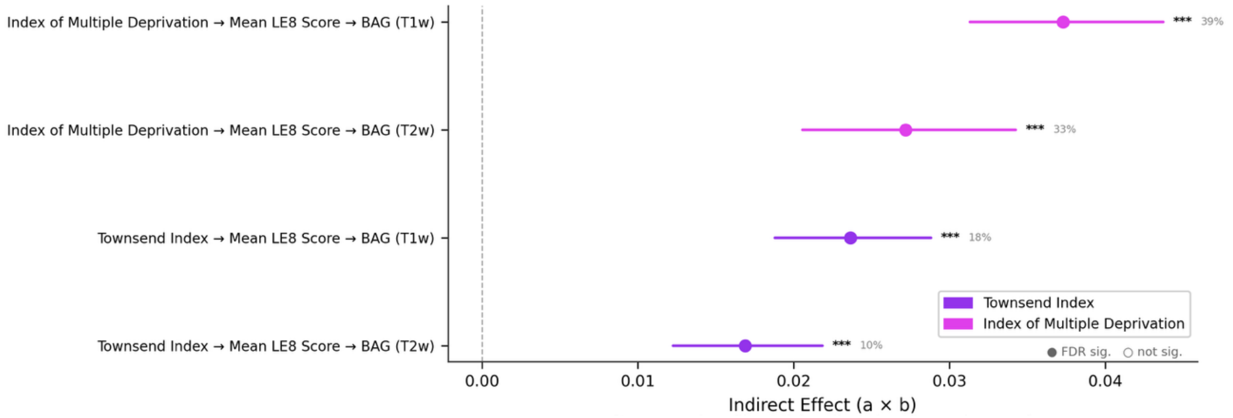


**eFigure 7 | LE8 mediates a greater proportion of the IMD-BAG association than the TDI-BAG association.** Forest plot of indirect effects (a x b) from the single mediator models testing the LE8 composite score as a mediator of the association between area-level deprivation (IMD and TDI) and BAG. Each row is one X → M → Y pathway; points are point estimates and horizontal bars are 5,000-iteration percentile bootstrap 95% confidence intervals. Pathways are colored by SES exposure (purple, TDI; pink, IMD). A positive indirect effect relates to lower SES being associated with higher BAG via worse LE8 score. Filled markers indicate pathways significant after Benjamini-Hochberg FDR correction (applied across all X~M~Y combinations in the run); open markers are non-significant. *** q < 0.001, ** *q* < 0.01, *q* < 0.05. Per-pathway proportion mediated is annotated to the right of each bar. Models adjusted for age, sex, and ethnicity (IMD n = 25,528; TDI n = 27,227). BAG, brain age gap; CI confidence interval; FDR, false discovery rate; Index of Multiple Deprivation, IMD; LE8, Life's Essential 8; SES, socioeconomic status; TDI, Townsend Deprivation Index

### eTable 5. Mediation by the Life's Essential 8 Score of Associations Between Socioeconomic Indicators and Brain Age Gap [abc]

| Exposure | Mediator | Outcome | No.[d] | Total effect, β (95% CI) | Total P | Path a (exposure→mediator), β (95% CI) | a P | Path b (mediator→outcome), β (95% CI) | b P | Direct effect, β (95% CI) | Direct P | Indirect effect, β (95% CI) | Indirect P (FDR) [e] | Proportion mediated |
|---|---|---|---|---|---|---|---|---|---|---|---|---|---|---|
| **Household income** | Life's Essential 8 score | Brain age gap (T1) | 25,011 | -0.085 (-0.123 to -0.048) | <.001 | 0.087 (0.076 to 0.098) | <.001 | -0.353 (-0.397 to -0.309) | <.001 | -0.054 (-0.092 to -0.017) | .004 | -0.031 (-0.036 to -0.025) | <.001 | 36.0% |
| **Townsend deprivation index** | Life's Essential 8 score | Brain age gap (T1) | 27,227 | 0.132 (0.093 to 0.171) | <.001 | -0.068 (-0.080 to -0.057) | <.001 | -0.347 (-0.389 to -0.305) | <.001 | 0.108 (0.069 to 0.148) | <.001 | 0.024 (0.019 to 0.029) | <.001 | 17.9% |
| **Household income** | Life's Essential 8 score | Brain age gap (T2) | 25,011 | -0.101 (-0.151 to -0.052) | <.001 | 0.087 (0.076 to 0.098) | <.001 | -0.266 (-0.324 to -0.207) | <.001 | -0.078 (-0.128 to -0.029) | .002 | -0.023 (-0.029 to -0.017) | <.001 | 22.7% |
| **Townsend deprivation index** | Life's Essential 8 score | Brain age gap (T2) | 27,227 | 0.170 (0.118 to 0.222) | <.001 | -0.068 (-0.080 to -0.057) | <.001 | -0.248 (-0.304 to -0.192) | <.001 | 0.153 (0.101 to 0.206) | <.001 | 0.017 (0.012 to 0.022) | <.001 | 9.9% |

Abbreviations: BAG, brain age gap; CI, confidence interval; FDR, false discovery rate; FLAIR, fluid-attenuated recovery; LE8, Life's Essential 8; TDI, Townsend Deprivation Index.

[a] Household income was modelled as an ordinal variable with higher values indicating higher income ; TDI was modelled continuously with higher values indicating greater area-level deprivation. Here, coefficients therefore carry opposite signs for each indictor whilst indicating the same direction of socioeconomic disadvantage.

[b] All effects are in years of brain age gap per 1-cateogry increase in income / 1-SD increase in TDI. Models were adjusted for age, sex, and ethnicity.

[c] Total, direct, and path coefficients were estimated by ordinary least squares Indirect effects and their CIs were estimated by percentile bootstrap with 5000 resamples.

[d] Sample sizes differ from those in eTabel 1 because analyses required participants with complete data for all LE8 components.

[e] P values were adjusted using the Benjamini-Hochberg false discovery rate across the 4 indirect effect tests reported in this table.

### eTable 6. Parallel Mediation by Individual Risk Factors of Associations Between Household Income and T1-weighted Brain Age Gap [a, b, c]

| Exposure | Mediator | Outcome | No. | Total effect, β (95% CI) | Total P | Path a (exposure→mediator), β (95% CI) | a P | Path b (mediator→outcome), β (95% CI) | b P | Direct effect, β (95% CI) | Direct P | Indirect effect, β (95% CI) | Indirect P (FDR) [d] | Proportion mediated |
|---|---|---|---|---|---|---|---|---|---|---|---|---|---|---|
| **Household income** | Diet risk score ↓(z) | Brain age gap (T1) | 16,886 | -0.089 (-0.135 to -0.043) | <.001 | -0.062 (-0.082 to -0.042) | <.001 | -0.014 (-0.050 to 0.022) | .45 | -0.051 (-0.097 to -0.005) | .03 | 0.001 (-0.001 to 0.003) | .57 | -1.0% |
| **Household income** | Pack years smoking (z) | Brain age gap (T1) | 16,886 | -0.089 (-0.135 to -0.043) | <.001 | -0.098 (-0.113 to -0.083) | <.001 | 0.223 (0.174 to 0.272) | <.001 | -0.051 (-0.097 to -0.005) | .03 | -0.022 (-0.028 to -0.016) | <.001 | 24.5% |
| **Household income** | Alcohol units weekly (z) | Brain age gap (T1) | 16,886 | -0.089 (-0.135 to -0.043) | <.001 | 0.035 (0.020 to 0.049) | <.001 | 0.366 (0.313 to 0.420) | <.001 | -0.051 (-0.097 to -0.005) | .03 | 0.013 (0.007 to 0.018) | <.001 | -14.2% |
| **Household income** | Physical inactivity (z) | Brain age gap (T1) | 16,886 | -0.089 (-0.135 to -0.043) | <.001 | 0.090 (0.076 to 0.104) | <.001 | -0.048 (-0.099 to 0.004) | .07 | -0.051 (-0.097 to -0.005) | .03 | -0.004 (-0.009 to 0.001) | .12 | 4.8% |
| **Household income** | Suboptimal sleep | Brain age gap (T1) | 16,886 | -0.089 (-0.135 to -0.043) | <.001 | -0.007 (-0.013 to -0.001) | .02 | 0.136 (0.017 to 0.255) | .02 | -0.051 (-0.097 to -0.005) | .03 | -0.001 (-0.002 to 0.000) | .10 | 1.0% |
| **Household income** | Systolic blood pressure (z) | Brain age gap (T1) | 16,886 | -0.089 (-0.135 to -0.043) | <.001 | -0.037 (-0.051 to -0.024) | <.001 | 0.249 (0.194 to 0.304) | <.001 | -0.051 (-0.097 to -0.005) | .03 | -0.009 (-0.013 to -0.006) | <.001 | 10.5% |
| **Household income** | HbA1c (%) (z) | Brain age gap (T1) | 16,886 | -0.089 (-0.135 to -0.043) | <.001 | -0.039 (-0.053 to -0.026) | <.001 | 0.233 (0.178 to 0.289) | <.001 | -0.051 (-0.097 to -0.005) | .03 | -0.009 (-0.013 to -0.006) | <.001 | 10.3% |
| **Household income** | BMI (z) | Brain age gap (T1) | 16,886 | -0.089 (-0.135 to -0.043) | <.001 | -0.048 (-0.062 to -0.035) | <.001 | 0.092 (0.038 to 0.146) | <.001 | -0.051 (-0.097 to -0.005) | .03 | -0.004 (-0.008 to -0.002) | .002 | 5.0% |

### eTable 6.Continued

| Exposure | Mediator | Outcome | No. | Total effect, β (95% CI) | Total P | Path a (exposure→mediator), β (95% CI) | a P | Path b (mediator→outcome), β (95% CI) | b P | Direct effect, β (95% CI) | Direct P | Indirect effect, β (95% CI) | Indirect P (FDR) [d] | Proportion mediated |
|---|---|---|---|---|---|---|---|---|---|---|---|---|---|---|
| **Household income** | Non-HDL Cholesterol (z) | Brain age gap (T1) | 16,886 | -0.089 (-0.135 to -0.043) | <.001 | 0.003 (-0.010 to 0.017) | .62 | -0.103 (-0.154 to -0.053) | <.001 | -0.051 (-0.097 to -0.005) | .03 | -0.000 (-0.002 to 0.001) | .71 | 0.4% |
| **Household income** | Infrequent social visits | Brain age gap (T1) | 16,886 | -0.089 (-0.135 to -0.043) | <.001 | 0.002 (-0.002 to 0.005) | .37 | -0.006 (-0.200 to 0.189) | .95 | -0.051 (-0.097 to -0.005) | .03 | -0.000 (-0.001 to 0.001) | >.99 | 0.0% |
| **Household income** | Stress (tense/restless) | Brain age gap (T1) | 16,886 | -0.089 (-0.135 to -0.043) | <.001 | -0.017 (-0.023 to -0.011) | <.001 | 0.060 (-0.055 to 0.175) | .30 | -0.051 (-0.097 to -0.005) | .03 | -0.001 (-0.003 to 0.001) | .41 | 1.1% |

Abbreviations: BAG, brain age gap; CI, confidence interval; FDR, false discovery rate; FLAIR, fluid-attenuated recovery; TDI, Townsend Deprivation Index.

[a] Household income was modelled as an ordinal variable with higher values indicating higher income

[b] All effects are in years of brain age gap per 1-cateogry increase in income Models were adjusted for age, sex, and ethnicity.

[c] Total, direct, and path coefficients were estimated by ordinary least squares Indirect effects and their CIs were estimated by percentile bootstrap with 5000 resamples.

[d] P values were adjusted using the Benjamini-Hochberg false discovery rate across the 4 indirect effect tests reported in this table.

### eTable 7. Parallel Mediation by Individual Risk Factors of Associations Between Household Income and T2-FLAIR Brain Age Gap [a, b, c]

| Exposure | Mediator | Outcome | No. | Total effect, β (95% CI) | Total P | Path a (exposure→mediator), β (95% CI) | a P | Path b (mediator→outcome), β (95% CI) | b P | Direct effect, β (95% CI) | Direct P | Indirect effect, β (95% CI) | Indirect P (FDR) [d] | Proportion mediated |
|---|---|---|---|---|---|---|---|---|---|---|---|---|---|---|
| **Household income** | Diet risk score ↓(z) | Brain age gap (T2) | 16,886 | -0.103 (-0.164 to -0.042) | <.001 | -0.062 (-0.082 to -0.042) | <.001 | -0.033 (-0.081 to 0.015) | .18 | -0.071 (-0.132 to -0.010) | .02 | 0.002 (-0.001 to 0.005) | .27 | -2.0% |
| **Household income** | Pack years smoking (z) | Brain age gap (T2) | 16,886 | -0.103 (-0.164 to -0.042) | <.001 | -0.098 (-0.113 to -0.083) | <.001 | 0.273 (0.207 to 0.339) | <.001 | -0.071 (-0.132 to -0.010) | .02 | -0.027 (-0.035 to -0.019) | <.001 | 26.0% |
| **Household income** | Alcohol units weekly (z) | Brain age gap (T2) | 16,886 | -0.103 (-0.164 to -0.042) | <.001 | 0.035 (0.020 to 0.049) | <.001 | 0.437 (0.367 to 0.506) | <.001 | -0.071 (-0.132 to -0.010) | .02 | 0.015 (0.008 to 0.022) | <.001 | -14.6% |
| **Household income** | Physical inactivity (z) | Brain age gap (T2) | 16,886 | -0.103 (-0.164 to -0.042) | <.001 | 0.090 (0.076 to 0.104) | <.001 | -0.015 (-0.085 to 0.056) | .69 | -0.071 (-0.132 to -0.010) | .02 | -0.001 (-0.008 to 0.005) | .77 | 1.3% |
| **Household income** | Suboptimal sleep | Brain age gap (T2) | 16,886 | -0.103 (-0.164 to -0.042) | <.001 | -0.007 (-0.013 to -0.001) | .02 | -0.025 (-0.183 to 0.132) | .75 | -0.071 (-0.132 to -0.010) | .02 | 0.000 (-0.001 to 0.001) | .77 | -0.2% |
| **Household income** | Systolic blood pressure (z) | Brain age gap (T2) | 16,886 | -0.103 (-0.164 to -0.042) | <.001 | -0.037 (-0.051 to -0.024) | <.001 | 0.141 (0.069 to 0.214) | <.001 | -0.071 (-0.132 to -0.010) | .02 | -0.005 (-0.009 to -0.002) | <.001 | 5.1% |
| **Household income** | HbA1c (%) (z) | Brain age gap (T2) | 16,886 | -0.103 (-0.164 to -0.042) | <.001 | -0.039 (-0.053 to -0.026) | <.001 | 0.251 (0.176 to 0.326) | <.001 | -0.071 (-0.132 to -0.010) | .02 | -0.010 (-0.015 to -0.006) | <.001 | 9.6% |
| **Household income** | BMI (z) | Brain age gap (T2) | 16,886 | -0.103 (-0.164 to -0.042) | <.001 | -0.048 (-0.062 to -0.035) | <.001 | 0.068 (-0.004 to 0.139) | .06 | -0.071 (-0.132 to -0.010) | .02 | -0.003 (-0.007 to 0.000) | .14 | 3.2% |
| **Household income** | Non-HDL Cholesterol (z) | Brain age gap (T2) | 16,886 | -0.103 (-0.164 to -0.042) | <.001 | 0.003 (-0.010 to 0.017) | .62 | -0.162 (-0.228 to -0.095) | <.001 | -0.071 (-0.132 to -0.010) | .02 | -0.001 (-0.003 to 0.002) | .77 | 0.5% |

### eTable 7. continued

| Exposure | Mediator | Outcome | No. | Total effect, β (95% CI) | Total P | Path a (exposure→ mediator), β (95% CI) | a P | Path b (mediator→outcome), β (95% CI) | b P | Direct effect, β (95% CI) | Direct P | Indirect effect, β (95% CI) | Indirect P (FDR) [d] | Proportion mediated |
|---|---|---|---|---|---|---|---|---|---|---|---|---|---|---|
| **Household income** | Infrequent social visits | Brain age gap (T2) | 16,886 | -0.103 (-0.164 to -0.042) | <.001 | 0.002 (-0.002 to 0.005) | .37 | 0.263 (0.007 to 0.519) | .04 | -0.071 (-0.132 to -0.010) | .02 | 0.000 (-0.001 to 0.002) | .55 | -0.4% |
| **Household income** | Stress (tense/restless) | Brain age gap (T2) | 16,886 | -0.103 (-0.164 to -0.042) | <.001 | -0.017 (-0.023 to -0.011) | <.001 | 0.142 (-0.012 to 0.297) | .07 | -0.071 (-0.132 to -0.010) | .02 | -0.002 (-0.005 to 0.000) | .14 | 2.3% |

Abbreviations: BAG, brain age gap; CI, confidence interval; FDR, false discovery rate; FLAIR, fluid-attenuated recovery; TDI, Townsend Deprivation Index.

[a] Household income was modelled as an ordinal variable with higher values indicating higher income

[b] All effects are in years of brain age gap per 1-cateogry increase in income Models were adjusted for age, sex, and ethnicity.

[c] Total, direct, and path coefficients were estimated by ordinary least squares Indirect effects and their CIs were estimated by percentile bootstrap with 5000 resamples.

[d] P values were adjusted using the Benjamini-Hochberg false discovery rate across the 4 indirect effect tests reported in this table

### eTable 8. Parallel Mediation by Individual Risk Factors of Associations Between Townsend Deprivation Index and T1-weighted Brain Age Gap [a, b, c]

| Exposure | Mediator | Outcome | No. | Total effect, β (95% CI) | Total P | Path a (exposure→mediator), β (95% CI) | a P | Path b (mediator→outcome), β (95% CI) | b P | Direct effect, β (95% CI) | Direct P | Indirect effect, β (95% CI) | Indirect P (FDR) [d] | Proportion mediated |
|---|---|---|---|---|---|---|---|---|---|---|---|---|---|---|
| **Townsend deprivation index** | Diet risk score ↓(z) | Brain age gap (T1) | 18,244 | 0.170 (0.120 to 0.220) | <.001 | 0.068 (0.046 to 0.090) | <.001 | -0.016 (-0.050 to 0.018) | .36 | 0.114 (0.064 to 0.163) | <.001 | -0.001 (-0.004 to 0.001) | .40 | -0.6% |
| **Townsend deprivation index** | Pack years smoking (z) | Brain age gap (T1) | 18,244 | 0.170 (0.120 to 0.220) | <.001 | 0.134 (0.118 to 0.151) | <.001 | 0.222 (0.175 to 0.269) | <.001 | 0.114 (0.064 to 0.163) | <.001 | 0.030 (0.023 to 0.037) | <.001 | 17.6% |
| **Townsend deprivation index** | Alcohol units weekly (z) | Brain age gap (T1) | 18,244 | 0.170 (0.120 to 0.220) | <.001 | 0.045 (0.029 to 0.060) | <.001 | 0.367 (0.315 to 0.419) | <.001 | 0.114 (0.064 to 0.163) | <.001 | 0.016 (0.011 to 0.023) | <.001 | 9.7% |
| **Townsend deprivation index** | Physical inactivity (z) | Brain age gap (T1) | 18,244 | 0.170 (0.120 to 0.220) | <.001 | -0.033 (-0.048 to -0.018) | <.001 | -0.042 (-0.091 to 0.006) | .09 | 0.114 (0.064 to 0.163) | <.001 | 0.001 (-0.000 to 0.003) | .13 | 0.8% |
| **Townsend deprivation index** | Suboptimal sleep | Brain age gap (T1) | 18,244 | 0.170 (0.120 to 0.220) | <.001 | 0.018 (0.011 to 0.024) | <.001 | 0.121 (0.006 to 0.236) | .04 | 0.114 (0.064 to 0.163) | <.001 | 0.002 (0.000 to 0.004) | .06 | 1.2% |
| **Townsend deprivation index** | Systolic blood pressure (z) | Brain age gap (T1) | 18,244 | 0.170 (0.120 to 0.220) | <.001 | -0.019 (-0.033 to -0.005) | .009 | 0.261 (0.208 to 0.313) | <.001 | 0.114 (0.064 to 0.163) | <.001 | -0.005 (-0.009 to -0.001) | .01 | -2.9% |
| **Townsend deprivation index** | HbA1c (%) (z) | Brain age gap (T1) | 18,244 | 0.170 (0.120 to 0.220) | <.001 | 0.026 (0.011 to 0.041) | <.001 | 0.248 (0.194 to 0.301) | <.001 | 0.114 (0.064 to 0.163) | <.001 | 0.006 (0.003 to 0.011) | <.001 | 3.8% |
| **Townsend deprivation index** | BMI (z) | Brain age gap (T1) | 18,244 | 0.170 (0.120 to 0.220) | <.001 | 0.041 (0.025 to 0.056) | <.001 | 0.073 (0.021 to 0.125) | .006 | 0.114 (0.064 to 0.163) | <.001 | 0.003 (0.001 to 0.006) | .01 | 1.7% |
| **Townsend deprivation index** | Non-HDL Cholesterol (z) | Brain age gap (T1) | 18,244 | 0.170 (0.120 to 0.220) | <.001 | -0.020 (-0.035 to -0.005) | .009 | -0.082 (-0.131 to -0.033) | .001 | 0.114 (0.064 to 0.163) | <.001 | 0.002 (0.000 to 0.003) | .02 | 1.0% |

### eTable 8. Continued.

| Exposure | Mediator | Outcome | No. | Total effect, β (95% CI) | Total P | Path a (exposure→ mediator), β (95% CI) | a P | Path b (mediator →outcome), β (95% CI) | b P | Direct effect, β (95% CI) | Direct P | Indirect effect, β (95% CI) | Indirect P (FDR) [d] | Proportion mediated |
|---|---|---|---|---|---|---|---|---|---|---|---|---|---|---|
| **Townsend deprivation index** | Infrequent social visits | Brain age gap (T1) | 18,244 | 0.170 (0.120 to 0.220) | <.001 | 0.005 (0.001 to 0.009) | .01 | -0.003 (-0.193 to 0.186) | .97 | 0.114 (0.064 to 0.163) | <.001 | -0.000 (-0.001 to 0.001) | .98 | -0.0% |
| **Townsend deprivation index** | Stress (tense/restless) | Brain age gap (T1) | 18,244 | 0.170 (0.120 to 0.220) | <.001 | 0.019 (0.013 to 0.026) | <.001 | 0.062 (-0.050 to 0.173) | .28 | 0.114 (0.064 to 0.163) | <.001 | 0.001 (-0.001 to 0.003) | .32 | 0.7% |

Abbreviations: BAG, brain age gap; CI, confidence interval; FDR, false discovery rate; FLAIR, fluid-attenuated recovery; TDI, Townsend Deprivation Index.

[a] TDI was modelled continuously with higher values indicating greater area-level deprivation.

[b] All effects are in years of brain age gap per 1-cateogry increase in income Models were adjusted for age, sex, and ethnicity.

[c] Total, direct, and path coefficients were estimated by ordinary least squares Indirect effects and their CIs were estimated by percentile bootstrap with 5000 resamples.

[d] P values were adjusted using the Benjamini-Hochberg false discovery rate across the 4 indirect effect tests reported in this table

### eTable 9. Parallel Mediation by Individual Risk Factors of Associations Between Townsend Deprivation Index and T2-FLAIR Brain Age Gap [a, b, c]

| Exposure | Mediator | Outcome | No. | Total effect, β (95% CI) | Total P | Path a (exposure→ mediator), β (95% CI) | a P | Path b (mediator →outcome ), β (95% CI) | b P | Direct effect, β (95% CI) | Direct P | Indirect effect, β (95% CI) | Indirect P (FDR) [d] | Proportion mediated |
|---|---|---|---|---|---|---|---|---|---|---|---|---|---|---|
| **Townsend deprivation index** | Diet risk score ↓(z) | Brain age gap (T2) | 18,244 | 0.230 (0.164 to 0.296) | <.001 | 0.068 (0.046 to 0.090) | <.001 | -0.031 (-0.078 to 0.015) | .18 | 0.164 (0.098 to 0.230) | <.001 | -0.002 (-0.006 to 0.001) | .24 | -0.9% |
| **Townsend deprivation index** | Pack years smoking (z) | Brain age gap (T2) | 18,244 | 0.230 (0.164 to 0.296) | <.001 | 0.134 (0.118 to 0.151) | <.001 | 0.271 (0.208 to 0.335) | <.001 | 0.164 (0.098 to 0.230) | <.001 | 0.036 (0.027 to 0.047) | <.001 | 15.9% |
| **Townsend deprivation index** | Alcohol units weekly (z) | Brain age gap (T2) | 18,244 | 0.230 (0.164 to 0.296) | <.001 | 0.045 (0.029 to 0.060) | <.001 | 0.436 (0.368 to 0.504) | <.001 | 0.164 (0.098 to 0.230) | <.001 | 0.019 (0.012 to 0.027) | <.001 | 8.5% |
| **Townsend deprivation index** | Physical inactivity (z) | Brain age gap (T2) | 18,244 | 0.230 (0.164 to 0.296) | <.001 | -0.033 (-0.048 to -0.018) | <.001 | 0.001 (-0.066 to 0.068) | .98 | 0.164 (0.098 to 0.230) | <.001 | -0.000 (-0.002 to 0.002) | >.99 | -0.0% |
| **Townsend deprivation index** | Suboptimal sleep | Brain age gap (T2) | 18,244 | 0.230 (0.164 to 0.296) | <.001 | 0.018 (0.011 to 0.024) | <.001 | -0.053 (-0.205 to 0.098) | .49 | 0.164 (0.098 to 0.230) | <.001 | -0.001 (-0.004 to 0.002) | .53 | -0.4% |
| **Townsend deprivation index** | Systolic blood pressure (z) | Brain age gap (T2) | 18,244 | 0.230 (0.164 to 0.296) | <.001 | -0.019 (-0.033 to -0.005) | .009 | 0.146 (0.076 to 0.215) | <.001 | 0.164 (0.098 to 0.230) | <.001 | -0.003 (-0.006 to -0.001) | .02 | -1.2% |
| **Townsend deprivation index** | HbA1c (%) (z) | Brain age gap (T2) | 18,244 | 0.230 (0.164 to 0.296) | <.001 | 0.026 (0.011 to 0.041) | <.001 | 0.266 (0.193 to 0.339) | <.001 | 0.164 (0.098 to 0.230) | <.001 | 0.007 (0.003 to 0.011) | <.001 | 3.0% |
| **Townsend deprivation index** | BMI (z) | Brain age gap (T2) | 18,244 | 0.230 (0.164 to 0.296) | <.001 | 0.041 (0.025 to 0.056) | <.001 | 0.047 (-0.022 to 0.116) | .19 | 0.164 (0.098 to 0.230) | <.001 | 0.002 (-0.001 to 0.005) | .24 | 0.8% |
| **Townsend deprivation index** | Non-HDL Cholesterol (z) | Brain age gap (T2) | 18,244 | 0.230 (0.164 to 0.296) | <.001 | -0.020 (-0.035 to -0.005) | .009 | -0.142 (-0.206 to -0.078) | <.001 | 0.164 (0.098 to 0.230) | <.001 | 0.003 (0.001 to 0.006) | .02 | 1.2% |

### eTable 9. continued

| Exposure | Mediator | Outcome | No. | Total effect, β (95% CI) | Total P | Path a (exposure→ mediator), β (95% CI) | a P | Path b (mediator →outcome ), β (95% CI) | b P | Direct effect, β (95% CI) | Direct P | Indirect effect, β (95% CI) | Indirect P (FDR) [d] | Proportion mediated |
|---|---|---|---|---|---|---|---|---|---|---|---|---|---|---|
| **Townsend deprivation index** | Infrequent social visits | Brain age gap (T2) | 18,244 | 0.230 (0.164 to 0.296) | <.001 | 0.005 (0.001 to 0.009) | .01 | 0.258 (0.009 to 0.507) | .04 | 0.164 (0.098 to 0.230) | <.001 | 0.001 (-0.000 to 0.003) | .10 | 0.6% |
| **Townsend deprivation index** | Stress (tense/restless) | Brain age gap (T2) | 18,244 | 0.230 (0.164 to 0.296) | <.001 | 0.019 (0.013 to 0.026) | <.001 | 0.138 (-0.012 to 0.287) | .07 | 0.164 (0.098 to 0.230) | <.001 | 0.003 (-0.000 to 0.006) | .10 | 1.1% |

Abbreviations: BAG, brain age gap; CI, confidence interval; FDR, false discovery rate; FLAIR, fluid-attenuated recovery; TDI, Townsend Deprivation Index.

[a] TDI was modelled continuously with higher values indicating greater area-level deprivation.

[b] All effects are in years of brain age gap per 1-cateogry increase in income Models were adjusted for age, sex, and ethnicity.

[c] Total, direct, and path coefficients were estimated by ordinary least squares Indirect effects and their CIs were estimated by percentile bootstrap with 5000 resamples.

[d] P values were adjusted using the Benjamini-Hochberg false discovery rate across the 4 indirect effect tests reported in this table

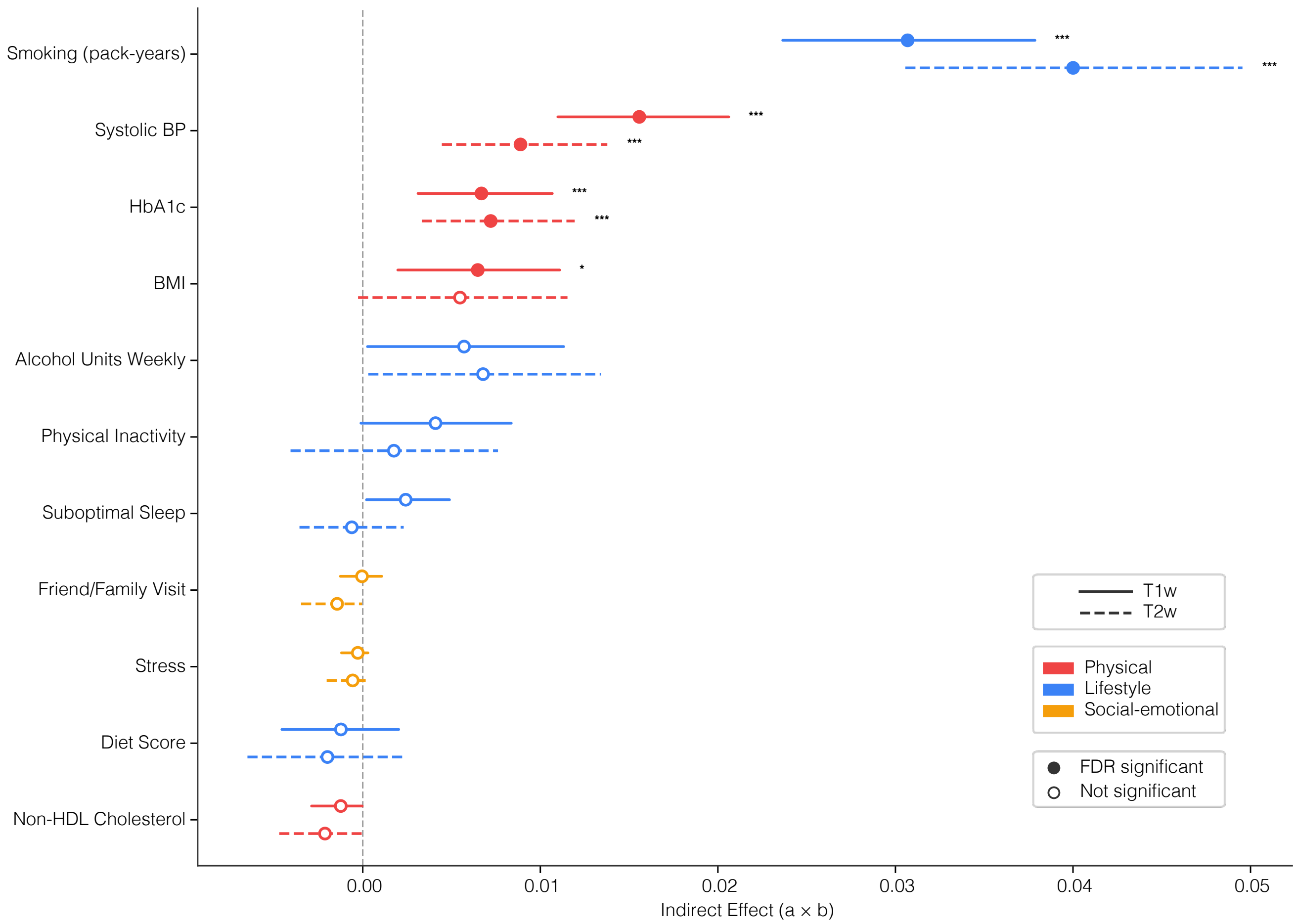


**eFigure 8 | Smoking carries most of the education→ BAG association.** Forest plot of specific indirect effects ($a_k x\ b_k$) from the parallel mediation model of education → BAG, with all eleven risk factors entered simultaneously as mediators. T1 BAG (solid) and T2 BAG (dashed) estimates are shown for each mediator. Each row is one mediator; points are point estimates and horizontal bars are 5000-iteration percentile bootstrap 95% confidence intervals. Mediators are colored by domain (red, physical; blue, lifestyle; yellow, social-emotional) and ordered within each panel by signed indirect effect (order row differs between panels). Education is coded as years of education, but the indirect effect has been sign flipped so that positive values describe pathways linking lower education to higher BAG (consistent with income and TDI figures). Filled markers indicate pathways significant after Benjamini-Hochberg FDR correction (applied across all X~M~Y combinations in the parallel mediation run); open markers are non-significant. *** $q < 0.001$, ** $q < 0.01$, $q < 0.05$. Per mediator proportion mediated is annotated to the right of each bar. For T2, all proportion mediated values are negative or near-zero because the of the opposite (protective) direction of effect for education-T2 BAG. Models adjusted for age, sex, and ethnicity (n=18,454). BAG, brain age gap; BMI, body mass index; BP, blood pressure; CI, confidence interval; FDR, false discovery rate.

### eReferences


1. Topiwala A, Ebmeier KP, Maullin-Sapey T, Nichols TE. Alcohol consumption and MRI markers of brain structure and function: Cohort study of 25,378 UK Biobank participants. *NeuroImage Clin*. 2022;35:103066. doi:10.1016/j.nicl.2022.103066

2. Petermann-Rocha F, Ho FK, Foster H, et al. Nonlinear Associations Between Cumulative Dietary Risk Factors and Cardiovascular Diseases, Cancer, and All-Cause Mortality: A Prospective Cohort Study From UK Biobank. *Mayo Clin Proc*. 2021;96(9):2418-2431. doi:10.1016/j.mayocp.2021.01.036

3. Singh SD, Oreskovic T, Carr S, et al. The predictive validity of a Brain Care Score for dementia and stroke: data from the UK Biobank cohort. *Front Neurol*. 2023;14:1291020. doi:10.3389/fneur.2023.1291020

4. Dibble A, Dalby C, Sevegnani M, et al. NeuroFM: Toward Precision Neuroimaging with Foundation Models for Individualized Brain Health Estimation. *medRxiv*. Published online April 1, 2026:2026.03.27.26349489. doi:10.64898/2026.03.27.26349489
